# Depiction of cell atlas and analysis of microenvironment alterations in human meniscus degeneration using single-cell transcriptomic

**DOI:** 10.1101/2022.05.19.22275311

**Authors:** Weili Fu, Sijie Chen, Runze Yang, Haoxiang Gao, Jian Li, Xuegong Zhang

## Abstract

**Background:** Musculoskeletal tissue degeneration impairs the life quality and function of many people. Meniscus degeneration is a major origin of knee osteoarthritis and a common threat to athletic ability, but its cellular mechanism remains elusive.

**Methods:** We built a cell atlas of healthy/degenerated human meniscus using scRNA-seq to investigate meniscal microenvironment homeostasis and its changes in the degeneration process and verified findings with immunofluorescent imaging.

**Results:** We identified and localized cell types in inner and outer meniscus, found new chondrocyte subtypes contributing to degeneration, and revealed how cellular compositions, functions, and interactions participated in degeneration.

We found that ECM disassembly, angiogenesis, and inflammation form a positive feedback loop driving the degeneration. Comparison of cellular interactions between different degenerative states identified each cell type’s functions in the loop.

**Conclusions:** The study revealed changes in the meniscal microenvironment during degeneration and discovered new cell subtypes as potential therapeutic targets. The observed mechanism could also be a general model for other joint degenerations.

**Funding:** The National Natural Science Foundation of China (81972123, 82172508, 62050178, 61721003) and the National Key Research and Development Program of China (2021YFF1200901).

## 1. Introduction

Degeneration of bones and joints are a major health issue for a large portion of the human population, especially for seniors, athletes and people of heavy physical labors. Aging and degeneration are complex processes characterized by the gradual loss of physiological functions over time. These are associated with increased susceptibility to diseases of many organ systems throughout the body. Aging is the strongest risk factor for degenerative bone and joint disease. The clinical treatment of age-related musculoskeletal disorders presents significant challenges because their pathogenic mechanisms remain unclear.

The knee is the largest and most complicated hinge joint associated with weight-bearing in the human body. The meniscus, an important component of the knee joint, plays essential roles in the load-bearing, shock absorption, nutrition, and lubrication of the knee joint articular cartilage and is very susceptible to injury and has a limited reparative potential^1–3^. Meniscal degeneration is one of the important risk factors for osteoarthritis (OA) and joint dysfunction, causing huge social and economic burdens^4^. It has been found through clinical observation that nearly half of patients with meniscal degeneration will eventually develop OA over time ^5^. Studies have proved that inflammatory responses and biomineralization affect the cellular microenvironment in the meniscus and contribute to the occurrence of disease^6^. However, the changes in the cell microenvironment in degenerated meniscus remain largely unclear. An adequate understanding of the changes that occur in the degenerative meniscus is critical for preventing meniscal injury in young and middle-aged patients and for relieving symptoms in elderly patients with knee OA. The meniscus can be anatomically divided into the inner region (the red zone) and the outer region (the white zone). Some researchers also use the “red-white” zone to denote the in-between transition region. There are blood vessels and nerves in the outer area, while few blood vessels and nerves are observed in the inner part. Aging and mechanical injuries may lead to meniscus degeneration. Different anatomical regions have varied recovery capabilities: the degeneration in the outer meniscus is more likely to heal while the degeneration in the inner part tends to be irreversible^7, 8^. The cell and molecular basis behind anatomical regions remain to be explored. It is unclear what cell type contributes to the inner and outer variations and what cell types mediate the degeneration.

The meniscal extracellular matrix (ECM) is the physics foundation of its biological roles, and alterations in the ECM may lead to meniscus degeneration and dysfunction. 72% of the meniscus wet weight is water, and the rest are the organic fractions (mainly cells and ECM) ^9^. Collagens account for the majority of the dry weight of the ECM. Different meniscal regions have different collagen compositions, and altered collagen composition is linked to meniscal disease. The outer meniscus is dominated by type II collagen, while the inner meniscus is dominated by type I collagen. Other kinds of collagens also existed in the meniscus (e.g., types III, IV, VI, XVIII)^10^. Proteoglycan (PG) is another glycosaminoglycans-containing macromolecule member of the meniscus ECM located within the collagen meshwork ^11^. The fragmentation of PG is a sign of meniscal degeneration. The major PG found in the meniscus is aggrecan^12^. The inner part of the meniscus has a relatively higher percentage of PGs compared with the outer^13^. Four kinds of glycosaminoglycans (GAGs) have been reported in the PG of healthy human meniscal tissue: chondroitin-6-sulfate, dermatan sulfate, chondroitin-4-sulfate, and keratin sulfate ^9, 14^. Some other non-collagen proteins (e.g., fibronectin, thrombospondin, tenascin) link the ECM to the meniscus cells and play a foundational role in this tissue ^15, 16^.

Building a detailed meniscal cell landscape is essential to understanding meniscus characteristics because the content of the extracellular matrix is synthesized and secreted by the meniscal cells. Early profiling of meniscal cell heterogeneities dates back to the work of Ghadially et al. in 1983^17^. They studied injured and uninjured human menisci with the electron microscope and stated that chondrocytes, a few fibroblasts, a few myofibroblasts, and a few intermediate state cells between chondrocytes and fibroblast existed in menisci. Scotti et al. also discussed that the cells in the superficial zone were fusiform while cells laid deeper were polygonal^18^. These classifications were mainly based on the shape and the surrounding matrix content. Some bulk RNA-seq studies^19^ revealed the up and down-regulated genes in the normal and degenerated tissues. Some studies proved the existence of stem/progenitor cells and senescent cells in cartilage tissue^20, 21^. Sun et al. identified the meniscus progenitors and reported their links to the progression of meniscal degeneration^22^. Following the knee articular cartilage single-cell cluster definitions provided by Ji et al.^23^, they identified cartilage progenitor cells (CPC), regulatory chondrocytes (RegC), prehypertrophic chondrocytes (PreHTC), hypertrophic chondrocytes (HTC), fibrochondrocytes (FC), fibrochondrocytes progenitors (FCP), and proliferating fibrochondrocytes (ProFC), degenerated progenitor (DegP), endothelial cells, and other immune cells. Their fine-granular cluster definitions covered many cell states and types in the meniscus. But their cluster markers may reflect some donor-specific molecular features and show limited generalization abilities for cell-type identification in new cell samples. Hence, a systematic profiling of cell types in both healthy and degenerated menisci is desirable.

With the recent development of single-cell omics, the researchers have investigated various cell types across multiple cartilage tissues, including knee joint cartilage^23^, intervertebral disc^24^, and meniscus^22^. These researches initiated a rudiment of the chondrocyte cell types and states, but the accumulated data covered an incomplete part of the heterogeneous cell subpopulations in the meniscus.

Our work established a single-cell transcriptomic atlas of 45,744 cells from 12 healthy/degenerated meniscus samples of 8 patients and provided an online cell browser at http://meni.singlecell.info:3000/ to support flexible explorations of the data. Our single-cell sequencing data has undergone rigorous quality control. Among the current single-cell sequencing studies involving articular cartilage and meniscus, we have the largest number of sequencing samples and cells. In addition, our data are the first single-cell sequencing study data involving inner and outer regions of the meniscus. We built a hierarchical cell type classification framework and defined five chondrocyte subtypes as well as two pericyte-like cell subtypes. Functional enrichment analysis revealed these subtypes’ roles in regulating ECM remodeling. We found that the loss of pericyte-like cells might facilitate vascularization in degenerated states. We profiled various immune cell types and observed damage-induced inflammatory responses in degenerated samples. Taken together, we inferred a positive feedback loop of three self-repairing processes, i.e., the ECM disassembly, inflammation, and angiogenesis, led to the chronic degeneration in the meniscal tissues. Once deviated from the homeostatic microenvironment, the three processes mutually reinforced each other and drove the development and progression of osteoarthritis. Meniscus is a representative fibrocartilage in the human body. The cellular and molecular changes during meniscal degeneration we observed may also be the mechanism of a variety of other cartilage degenerations. The discovery may suggest new ideas for the treatment of all types of joint degeneration in the body.

## 2. Results

### 2.1. The cellular landscape of the inner and outer parts of human menisci

To understand the composition and molecular profiles of the cells from different regions and degeneration status, we randomly sampled four healthy menisci and four degenerated menisci from the patient cohort and performed single-cell RNA sequencing. Figure 1A illustrates the morphological patterns of the samples: meniscal specimens in the normal group are smooth and complete with a clear structure. The specimens from the degenerated group are swollen, irregular in shape, and damaged in structure. Each specimen’s inner part (white zone) and the outer part (red zone) were strictly separated and collected. These samples are next dissociated and prepared for single-cell RNA sequencing (Methods) as shown in the overall workflow (Figure 1B).

**Figure 1:**
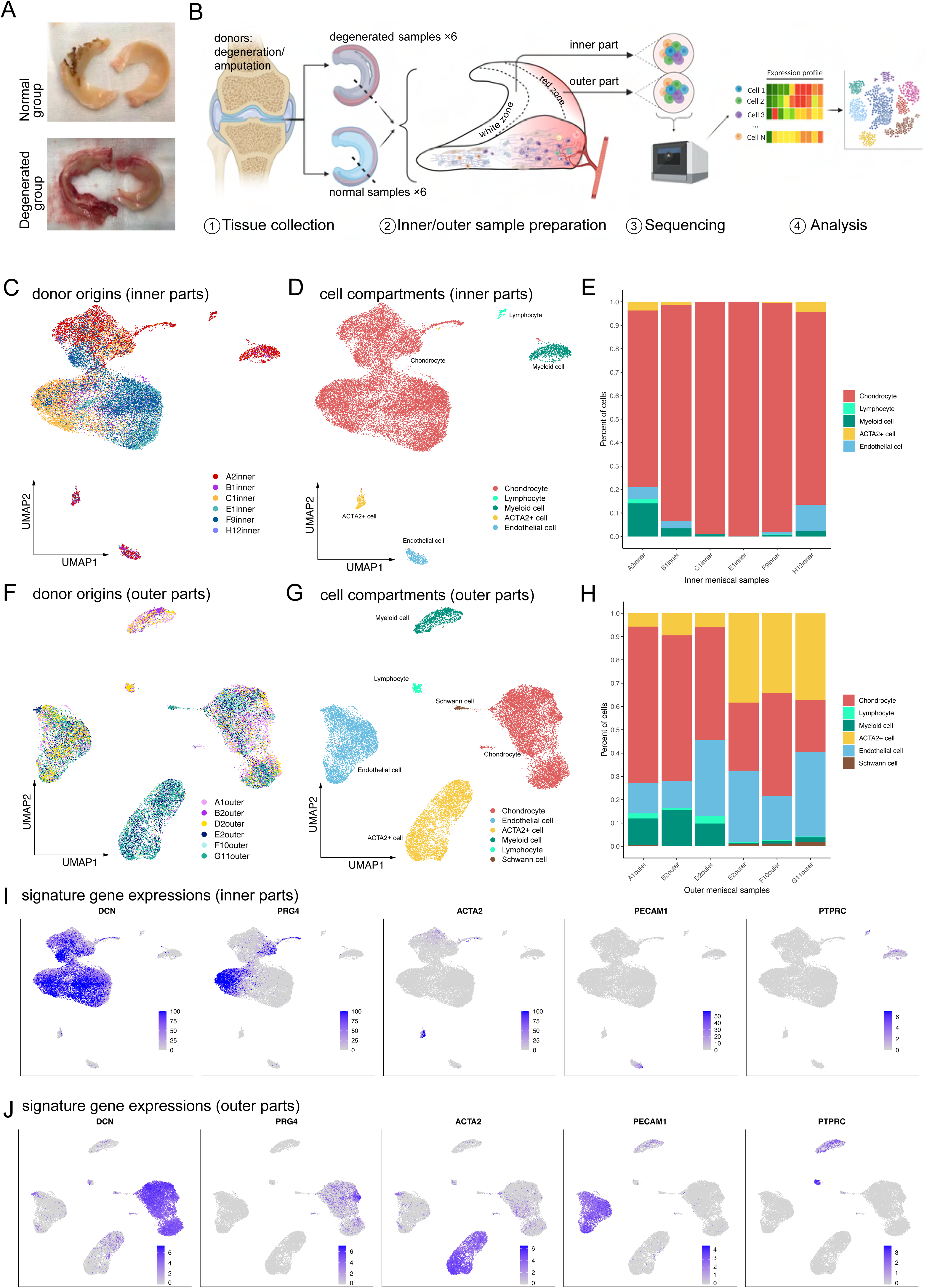
Single-cell RNA-seq reveals major cell classes in human menisci. (A) Photographs of typical normal and degenerative meniscus specimens. B: Meniscal cells from the inner parts and the outer parts of donor menisci are sent for single-cell RNA sequencing library construction. (B) The overall workflow of the single-cell sequencing. Inner and outer meniscal parts are collected separately from patients with normal/degenerated menisci. (C), (F) UMAP visualization of the donor origins in inner/outer samples. (D), (G) UMAP visualization of the major cell classes. In the coming contents, we show fine-grained clusters under the classes. The cell classes were determined based on known marker genes. (E) The percentages of the identified cell classes in six samples of the inner meniscus. (H) The percentages of the classes in six samples of the outer meniscus. (I), (J): The expression levels of the major class markers of the inner cells (up) and the outer cells (down). Darker colors indicate higher expression levels.

After comprehensive quality control steps removing low-quality cells and inferred doublets (Methods), we recovered 45,744 cells from 12 samples of 8 people. The number of counts observed per cell and the number of genes observed per cell are visualized in Supplementary Figures S1 A-B. We can infer from the quality control steps that the general sequencing quality is good except for the E2outer and H12inner samples.

Here we show the inner and outer menisci’s major cell classes. An unsupervised clustering algorithm partitioned the cells into separated clusters (Methods), which were next visualized using uniform manifold approximation and projection (UMAP). UMAP and stacked bar plots in Figure 1 C-E described the distributions of cells in the inner menisci, and Figure 1 F-H described them in the outer menisci. Figure 1C and Figure 1F visualized the origin of the cells, indicating that the batch effects of the samples were well eliminated, and the biological variations were preserved. The cell type correlation plots in Supplementary Figure 1C also confirmed that cells of the same type, not the same batch, had higher affinities. We observed five major cell classes in the inner menisci: chondrocytes, lymphocytes, myeloid cells, endothelial cells, and a group of ACTA2^+^ cells. We also observed the five classes in the outer menisci, plus a group of Schwann cells, indicating there are nerve tissues in the outer menisci.

The percentages of cells in Figure 1E and Figure 1H showed that, in general, the outer menisci had higher cell type diversities. In the more diverse outer menisci, the degenerated group (A1, B2, D2) had higher endothelial and immune cells ratios and lower ACTA2+ cell ratios.

The cell classes were determined by combinations of markers in Figure 1 I-J and Supplementary Figure 2 A-B. We identified cell groups as chondrocytes if they expressed a high level of cartilage-related genes such as DCN, PRG4, COL1A2, COL3A1, ACAN, COMP, FN1, CHI3L1, and CHI3L2. We identified cell groups with high expression levels of PLVAP, PECAM1 as endothelial cells. Immune cells were identified using high PTPRC, myeloid markers CD68, C1QA, HLA-DQA1, and T cell markers CD3D, CD3E. In addition, there are a group of cells separated from other cells in both inner and outer areas of the meniscus, which has high expression levels of ACTA2. Besides high expression of ACTA2, this population of cells also expressed some chondrocyte-associated gene markers, such as COL1A2, COL3A1, DCN, and FN1. We tentatively named this ACTA2^+^ population pericyte-like cells or PCL (Figure 1I, J and Supplementary Figure 2A, B) and reanalyzed them in the PCL class together with the chondral class in Figure 2.

**Figure 2:**
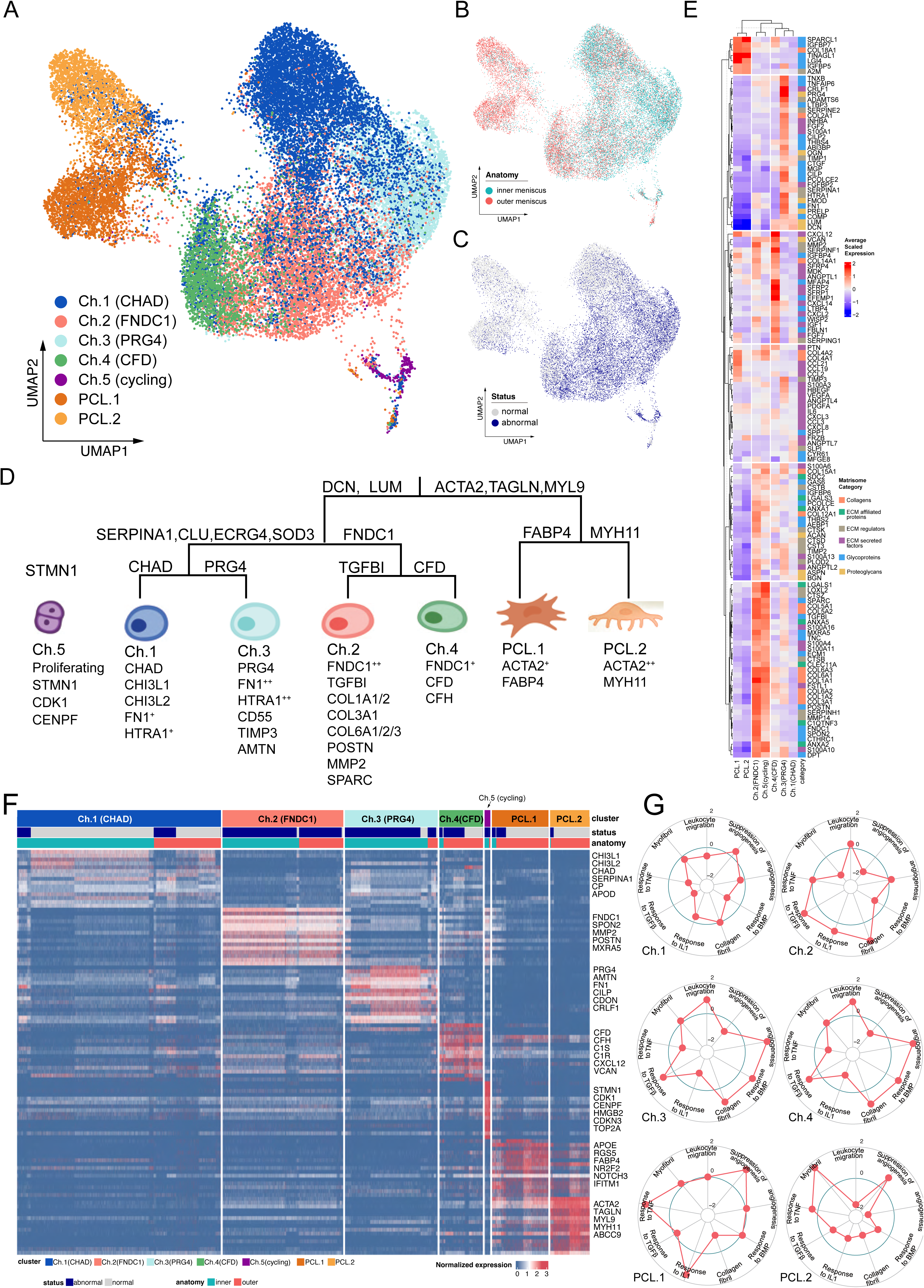
Identification of chondrocyte and PCL subclusters in human meniscus. (A) UMAP visualization of the chondrocyte and PCL class cells. (B) and (C) UMAP visualization of the distribution of chondrocytes at different anatomical sites (up) and different sample statuses (down). (D): A schematic diagram of the hierarchical classification of chondrocyte subgroups. The classification criteria are along the tree path, and each group’s highly expressed marker genes are given below each cluster. (E) A cluster-level heatmap shows ECM-related gene expressions in each chondrocyte/PCL subpopulation. (F) A cell-level heatmap reveals the normalized expression of differentially expressed genes for each cluster defined in (D). (G) Radar plots score nine molecular themes of each cell type (leukocyte migration, suppression of angiogenesis, angiogenesis, response to BMP, collagen fibril, response to IL1, response to TGFβ, response to TNF, and myofibril). GSVA calculated the scores using themes picked from MSigDB.

### 2.2. Functional coordination of the heterogeneous chondrocytes and PCL cells

We built a comprehensive cell identity framework charting the meniscal chondrocytes and PCL cells into a hierarchical coordination system. Unlike the other single-cell cartilage studies that assigned cell types with several marker genes, we defined cell types in a top-down way to simultaneously reveal subtle granular-view variations and exhibit general similarities.

To propose such a cell identity framework, we took multiple factors into account: clustering labels, clustering granularities, marker gene expression specificity, generalization abilities across samples and conditions, supporting evidence in other studies, matrix compositions, and biological functions (Methods). We finally identified five chondral clusters named chondrocytes-1 to chondrocytes-5 (Ch.1 ∼ Ch.5 in Figure 2A) and two pericyte-like cell clusters (PCL.1 and PCL.2 in Figure 2A) which were named as ACTA2^+^ cells in Figure 1D and Figure 1G. The distribution shown in figure 2B and C and the correlations in Supplementary Figure S4A indicated that the batch effects are resolved well. As described in Figure 1E/H, PCL.1 and PCL.2 are enriched in the normal samples, and Ch.2 and Ch.3 are enriched in the degenerated samples (Figure 2C).

We visualized the cell type definition framework in a tree in Figure 2D, which assigned labels to cells with hierarchical marker combinations. Common fibroblast markers like DCN, LUM are highly expressed in the chondral class. Smooth muscle cells markers like ACTA2, TAGLN, MYL9 are highly expressed in the pericyte-like cell (PCL) class. More detailed chondrocyte subpopulation markers are visualized in Figure 2F, Supplementary Figure S4B, and tested in Supplementary Figure S9B and S9C. We further demonstrated that these subclusters played multifaceted roles in meniscal homeostasis and degeneration progression under the branches (Figure 2G, Supplementary Figure S3).

For the granular cell subtypes, we analyzed their biological functions using gene set overrepresentation enrichment analysis ^25^ and gene set variation analysis ^26^ (Figure 3C-F and Supplementary Figure S3A-S3G). The functional analysis identified subtypes that contributed to the angiogenesis/anti-angiogenesis process, and subtypes that contributed to the ECM construction/disassembly process. See coming sections for detailed subtypes and functions.

**Figure 3:**
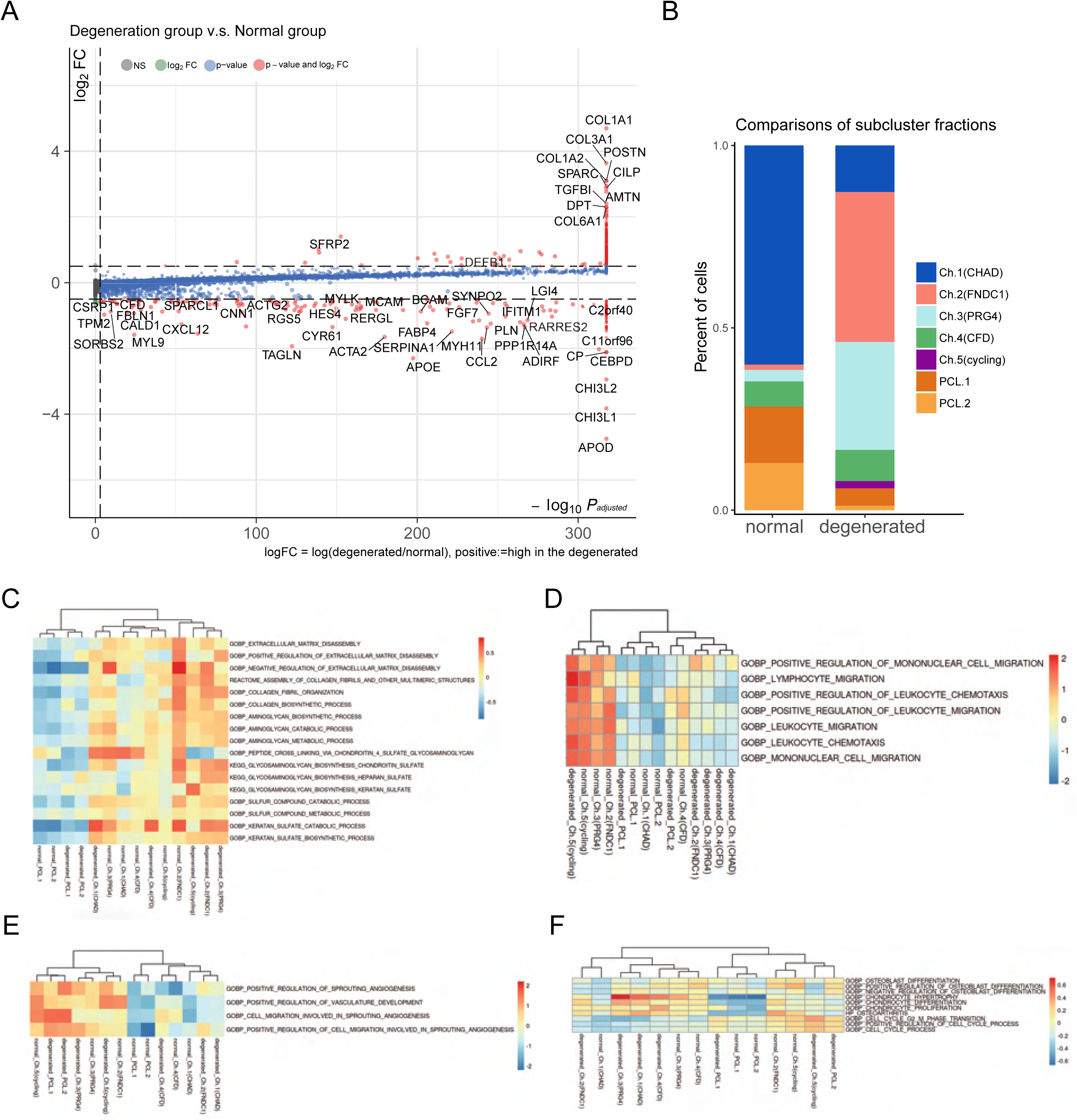
Degeneration molecular patterns in chondrocytes and PCLs. (A) A volcano plot shows differentially expressed genes with high fold change values. The comparison was made between 1) chondrocytes and PCLs in the degenerated group; 2) chondrocytes and PCLs in the normal group. (B) The proportion of each subcluster of chondrocytes in the degenerated and normal meniscus. (C)-(F) Gene set variation analysis of each cluster under degenerated/normal conditions. Gene sets supported functions are evaluated using GSVA (Methods), and scaled values are visualized in the heatmaps. We observed Ch.2, Ch.3 had high scores for matrix disassembly activities, angiogenesis activities, chemotaxis activities, and chondrocyte hypertrophy regardless of the normal/degeneration status. The cycling chondrocytes Ch.5 have high scores in the cell-cycle related terms.

We constructed gene regulatory networks (GRN) using pySCENIC ^27, 28^ to decipher revealed the common and unique programs behind the chondrocyte subpopulations and the PCL populations (Methods). In the inferred gene regulatory network, links originate from the upstream TFs to the downstream targets. Each TF forms a module of nodes that contains its targets. We calculated the module activation scores and showed the binarized status of the gene regulatory programs in Supplementary Figure S8A. Supplementary Figure S8B visualizes the core part of the network. We found that chondrocyte subcluster Ch 2-5 shared the common gene regulatory modules TRPS1, FOXC1, and HOXD10, but each of them possessed some specific modules, for example, Ch.2 had AKR1A1, Ch.3 had SIX3, Ch.4 had CEBPA, and Ch.5 had MYBL2. This GRN analysis also indicated that though sharing some transcriptomic features of chondrocytes, PCL cells were a distinct population with different gene regulatory programs. The GRN analysis also explained the heterogeneity of the two subpopulations inside PCL cells.

### 2.3. Pericyte-like cells stabilize the blood vessels in the meniscus microenvironment

In the pericyte-like cell class, the cells present strong pericyte/muscle cell signature genes like ACTA2 and MYL9 (Figure 2F, Supplementary Figure S2) and appear more frequently in the normal outer menisci (Figure 2C, Figure 2F). The marker gene enrichment analysis reported enriched GO terms such as “muscle contraction”, “muscle organ development”, “smooth muscle cell proliferation” in these cells (Supplementary Figure S3F). Our Immunofluorescence staining revealed that this population of cells lay around the vascular endothelial cells and formed a cube shape outside the vessels (Figure 4A-4B). Hence, we decided to name them pericyte-like cells (PCL).

**Figure 4:**
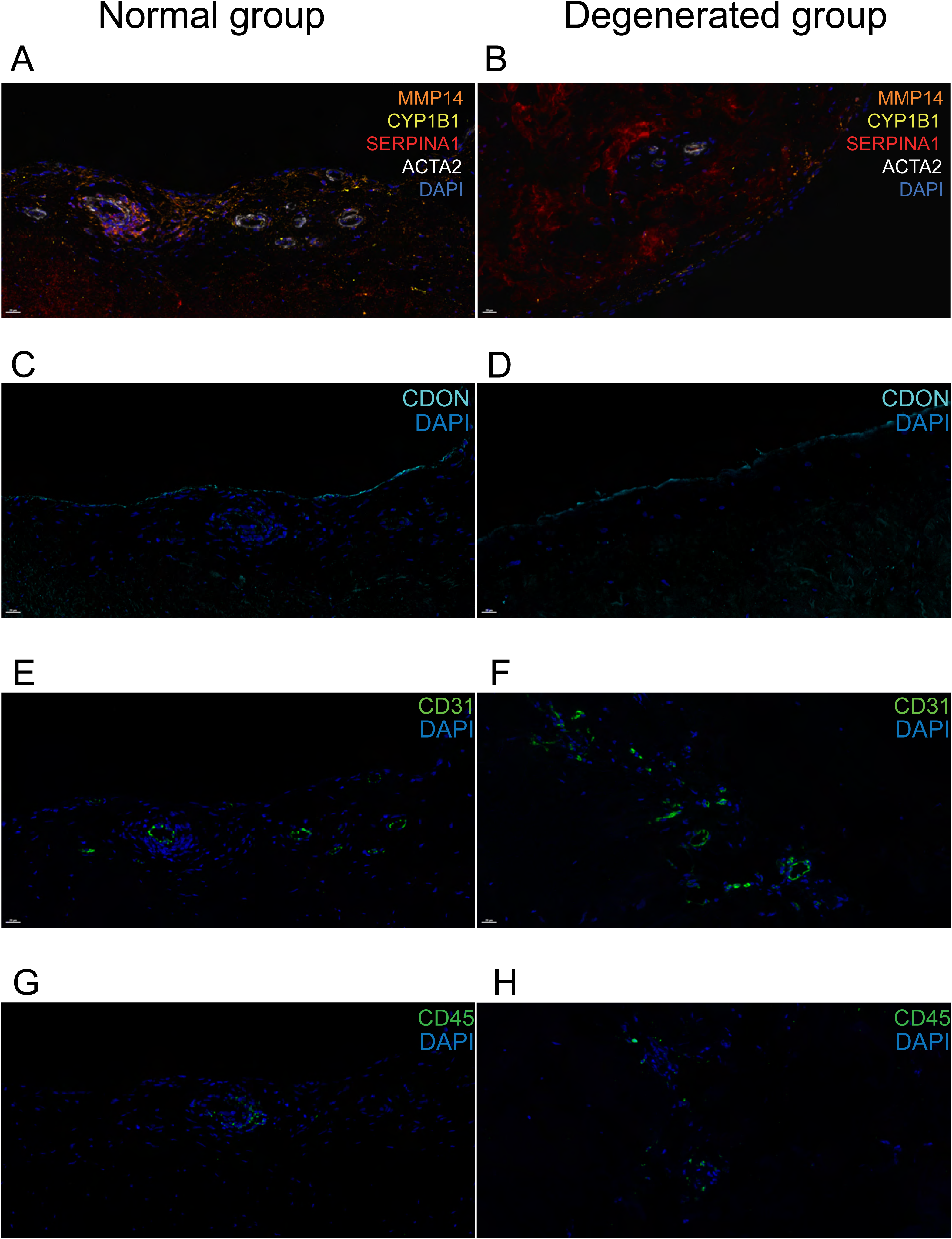
Immunofluorescent staining of human menisci demonstrating resident cell subtypes. Representative immunofluorescence staining images. Color settings: MMP14 for Ch.2 cells (orange), SERPINA1 for Ch.1 cells (magenta), ACTA2 for PCL cells (white), CDON for Ch.3 cells (cyan), CYP1B1 for Ch.4 cells (yellow), CD31 for endothelial cells (green), CD45 for immune cells (green). Nuclei are stained blue (DAPI). Scale bar 20 μm. (A) Multiplexed IF staining of the normal meniscus. (B) Multiplex IF staining of the degenerated meniscus. (C) IF staining of CDON in a normal sample. (D) IF staining of CDON in a degenerated sample. (E) IF staining of CD31 in a normal sample. (F) IF staining of CD31 in a degenerated sample. (G) IF staining of CD45 in a normal sample. (H) IF staining of CD45 in a degenerated sample. (IF: Immunofluorescence)

We identified two subtle subclusters within the PCL class. One subcluster PCL.1 expresses a higher level of FABP4, RGS5, and the other PCL.2 subcluster expresses a higher level of MYH11, MYH9, CNN1, TAGLN. Kumar, et al. ^29^ has analyzed these genes’ functions in the specification and diversification of pericyte/smooth muscle cells from mesenchymoangioblasts.

Hence, it is likely that PCL.1 has the properties of type 2 pericyte in their study, and PCL.2 has the properties of the smooth muscle cells. GSVA gene set activity analysis revealed that PCL.1 had more active responses to IL1/TNF, indicating this subcluster may participate in the inflammatory responses, while PCL.2 had higher myofibril gene set activation scores. Both subclusters suppress the angiogenesis process. From the ECM perspective, average ECM-related gene expression analysis showed that PCL synthesis ECM glycoproteins encoded by SPRCL1, IGFBP7, TINAGL1, LGI4, IGFBP5. Antiangiogenic genes like COL4A1, COL4A2, COL18A1 are highly expressed in PCL.1 (Supplementary Figure S12B). Secreted chemokines like CCL2, CCL19, CCL21, and CXCL12 indicated PCL.1 might have essential rules in recruiting lymphoid cells like T cells (Supplementary Figure S12D).

Though there are articles reporting cells similar to PCLs, we did not directly adopt their notations. Gan, et al. ^24^ named a group of ACTA2^+^ cells as pericytes in a single-cell study profiling human intervertebral discs. “Pericyte-like cells” would be a more rigorous term in our work because we can only prove these populations have some pericyte/SMC-like phenotypes. In the work of Sun, et al. ^22^, they named a similar ACTA2^+^ MYLK^+^ MCAM^+^ MYL9^+^ cluster as the fibrochondrocytes progenitor (FCP), claimed they differentiated into degenerated meniscus progenitors (DegP) in degenerated samples, and differentiate into various lineages if ex vivo cultured. However, our observations did not find clustered cells expressing DegP markers GREM1, CDCP1, DNER (Supplementary Figure S10A). The cells expressing these markers are scattered and not topologically connected with the ACTA2^+^ MYLK^+^ MCAM^+^ MYL9^+^ in our observations (Figure 2A, Supplementary Figure S10A, and Fig4B in Sun, et al. ^22^), which weaken the evidence that our PCLs differentiate into DegP in our data.

It is known that pericytes adhere to the surface of endothelial cells ^30, 31^, which are building blocks of blood vessels, maintain a stable vascular network via endothelial-pericyte crosstalk. We inferred pericyte-like cells in meniscal microenvironments played similar anti-angiogenesis roles like the pericytes according to the gene set variation analysis (Figure 2G) and the decreased percentages of PCLs (Figure 2F) in the degeneration group.

### 2.4. Chondrocyte subpopulations and their roles in ECM remodeling

The extracellular matrix (ECM) in meniscal tissues is continuously undergoing remodeling processes to keep homeostasis of the microenvironment. We aim to explain the roles of chondrocyte subpopulations that synthesize the ECM in the remodeling process in different degeneration states.

With the hierarchical cell type definition framework, we identified two major branches and a small cluster undergoing cell cycle phase transitions in the chondrocyte class. These branches of cells have varied roles in the construction and maintenance of the extracellular matrix, angiogenesis, and chemotaxis of immune cells.

The largest branch is the chondrocyte-1 (Ch.1) population that highly expresses CHAD, CHI3L1, CHI3L2, ECRG (C2orf40), and APOD. Some of these genes (e.g., CHI3L1/2) have been described as the markers of regulatory chondrocytes (RegC) ^23^. This cluster’s percentage decreased in the degeneration group. The gene set activation analysis indicated that this population of cells negatively regulates the angiogenesis process. In Figure 2E, we visualized the gene expressions that play essential roles in the extracellular matrix according to the Matrisome project ^32^. ECM-related gene expressions indicated Ch.1 produces ECM regulators TIMP1, TIMP4, which were inhibitors of ECM destruction proteins. Hence, we inferred that the CHAD^+^ Ch.1 population is a cluster of chondrocytes that maintain homeostasis in the meniscal tissue.

One branch that highly expresses FNDC1 and THY1 has two subclusters. One TGFBI^+^ subcluster has particularly high expression levels of FNDC1, so we named it chondrocyte2-FNDC1 (Ch.2). Ch.2 highly expresses type I, III, VI collagen (COL1A1/2, COL3A1, COL6A1/2/3), POSTN, MMP2, SPARC, and MXRA5, which are reported as osteoarthritis marker genes in previous studies^19^ (Supplementary Figure S9 A), and are overlapped with part of the chondrocyte hypertrophy genes^23^. Matrix remodeling-associated protein 5 (MXRA5) is a TGF-β1 regulated protein, which is consistent with our observation that Ch.2 has a high gene set activity score of “Response to TGFβ” in Figure 2G. FNDC1 and MXRA5 have been recently reported as new markers of aortic stenosis, which were involved in the chondrocyte-mediated valve calcification ^33^. Apart from MMP2, this population also expresses matrix destruction proteins like MMP11/13/14, ADAM9/12, ADAMTS2 (Supplementary Figure S12 A). With all information considered, we inferred that Ch.2 (FNDC1) is a chondrocyte population that is associated with aberrant extracellular matrix remodeling, and this population may contribute to the progress of human meniscal degeneration. We named the CFD^+^ subcluster expressing high levels of CFD, CFH, C1R, C1S, CXCL12, VCAN as Ch.4 (CFD). This subcluster expresses genes associated with alternative complement activation pathways (C1R, C1S, C2, C3, C6, C7) and may promote inflammation by attracting macrophages and neutrophils (Supplementary Figure S12 C).

We named another branch that expresses high levels of PRG4, HTRA1, FN1, CD55, TIMP3, CDON, AMTN as Ch.3 (PRG4). We inferred that Ch.3 plays a lubrication role on the surface of the meniscus and also appears in the degeneration reformed region because proteoglycan 4 (PRG4) has a lubrification function within the articular cartilage surface. Our immunofluorescence staining further confirmed that CDON expresses along the surface region of the meniscus (Figure 4). This population highly expresses TIMP1/2/3 (Supplementary Figure S12 A) and may balance the MMP/ADAM/ADAMTS producing cells’ production and inhibit the matrix decomposition.

We did not directly adopt the chondrocyte terminology used in other researches such as Ji, et al. ^23^ and Sun, et al. ^22^, and instead used Ch.1 to Ch.5 to represent the subclusters because our data spans chondrocytes of higher quantity and greater heterogeneity. We have examined some literature-derived markers on our data in Supplementary Figures S10-S11, which produced less clear or contradictory label inferences.

### 2.5. Degeneration associated cell subpopulation changes

We compared the overall differences between the normal chondrocytes and the degenerated chondrocytes using differentially expressed genes (DEG) analysis. We identified genes upregulated in the degeneration group such as COL1A1, COL1A2, COL3A1, COL6A1, POSTN, SPARC, CILP, TGFBI, DPT, ATMN and genes upregulated in the normal group such as APOD, CHI3L1, CHI3L2, CEBPD, C11orf96, C2orf40, ACTA2, TAGLN, BCAM, MCAM (Figure 3A, extended gene list in Supplementary Figure S5, S6A). Functional interpretation of these marker genes is available in Supplementary Figure S6 B-C. We believe these differential expressions are mainly induced by the cell type compositional changes because these genes are identified as markers of the chondrocyte subpopulations. For example, the Ch.2 marker genes COL1A1/2, COL3A1, COL6A1, SPARC are highly expressed in the degenerated group; the Ch.3 markers AMTN, CILP are also highly expressed in the degenerated group; the markers of the pericyte-like cells, ACTA2, TAGLN, BCAM, MCAM are highly expressed in the normal group. In addition, we observed an increase in the percentage of Ch.2/3 and a decrease in that of Ch.1 and PCL (Figure 3B and Supplementary Figure S7A), which were consistent with the DEG analysis. We also compared the DEGs in each chondrocyte subpopulation between the degenerated and normal groups (Supplementary Figure S7B) and found that the DEGs in each subcluster were different from overall DEGs (Supplementary Figure S5). All the above factors considered, we concluded that the expressional changes of the meniscus during meniscus degeneration were constitutively contributed by the changes in the proportion of meniscal cell subpopulations.

The cluster-level gene set variation analysis (Figure 3C) further confirmed that Ch.2 and Ch.3 played essential roles in the anabolism/catabolism balance of the ECM regardless of the normal/degenerated status. Ch.2 and Ch.3 also got high scores in chemotaxis and angiogenesis-related terms in normal and degenerated groups (Figure 3D-E). Ch.1 and normal group PCL got low scores in the angiogenesis-related terms. Ch.5 are scored high in the cell-cycle-related terms (Figure 3F). These gene set activity variations also prompted that the subclusters’ biological functions are relatively stable across the normal/degenerated conditions, and the subclusters’ percentage change contributed to the degeneration process.

### 2.6. Spatially resolve cell subtype distributions with in situ imaging

We used immunofluorescence staining to validate the subpopulation markers and study the spatial distribution of the subclusters in the human meniscus. Among the top 20 marker genes (log2 fold change values) for each chondrocyte subpopulation, we chose SERPINA1, MMP14, CDON, CYP1B1, ACTA2, CD31, and CD45 for staining. Normal and degenerated meniscus sample slides were simultaneously stained with the following antibodies: anti-SERPINA1 for Ch1(CHAD), anti-MMP14 for Ch2(FNDC1), anti-CYPIB1 for Ch4(CFD), and anti-ACTA2 for PCL cells. We used anti-CD31, anti-CDON, and anti-CD45 to label vascular endothelial cells, Ch3(PRG4), and immune cells for immunofluorescence staining. From the staining results, we can clearly find that CDON+ chondrocytes were mainly distributed on the surface of the meniscus tissue, and ACTA2^+^ chondrocytes were presented surrounding blood vessels.

SERPINA1^+^, CYP1B1^+,^ and MMP14+ chondrocytes were mixed and scattered inside the meniscus. Immune cells were mainly distributed around the blood vessels in the meniscus. And comparing with the normal samples, the number of ACTA2^+^ chondrocytes were decreased, the number of MMP14+ chondrocytes was markedly increased, and the number of vascular endothelial cells were significantly increased in degenerative specimens (Figure 4A-H).

### 2.7. Endothelial cells, immune cells and chondrocytes form a positive feedback loop of degeneration

Besides chondrocytes, endothelial cells and various types of immune cells also have indispensable roles in the occurrence and development of cartilage degeneration and osteoarthritis ^34, 35^.

In our meniscal samples, we identified macrophages/monocytes, neutrophils, mast cells, dendritic cells, T cells, and cycling immune cells that are hard to recognize identities (Figure 5A-B, Supplementary Figure S14A).

**Figure 5:**
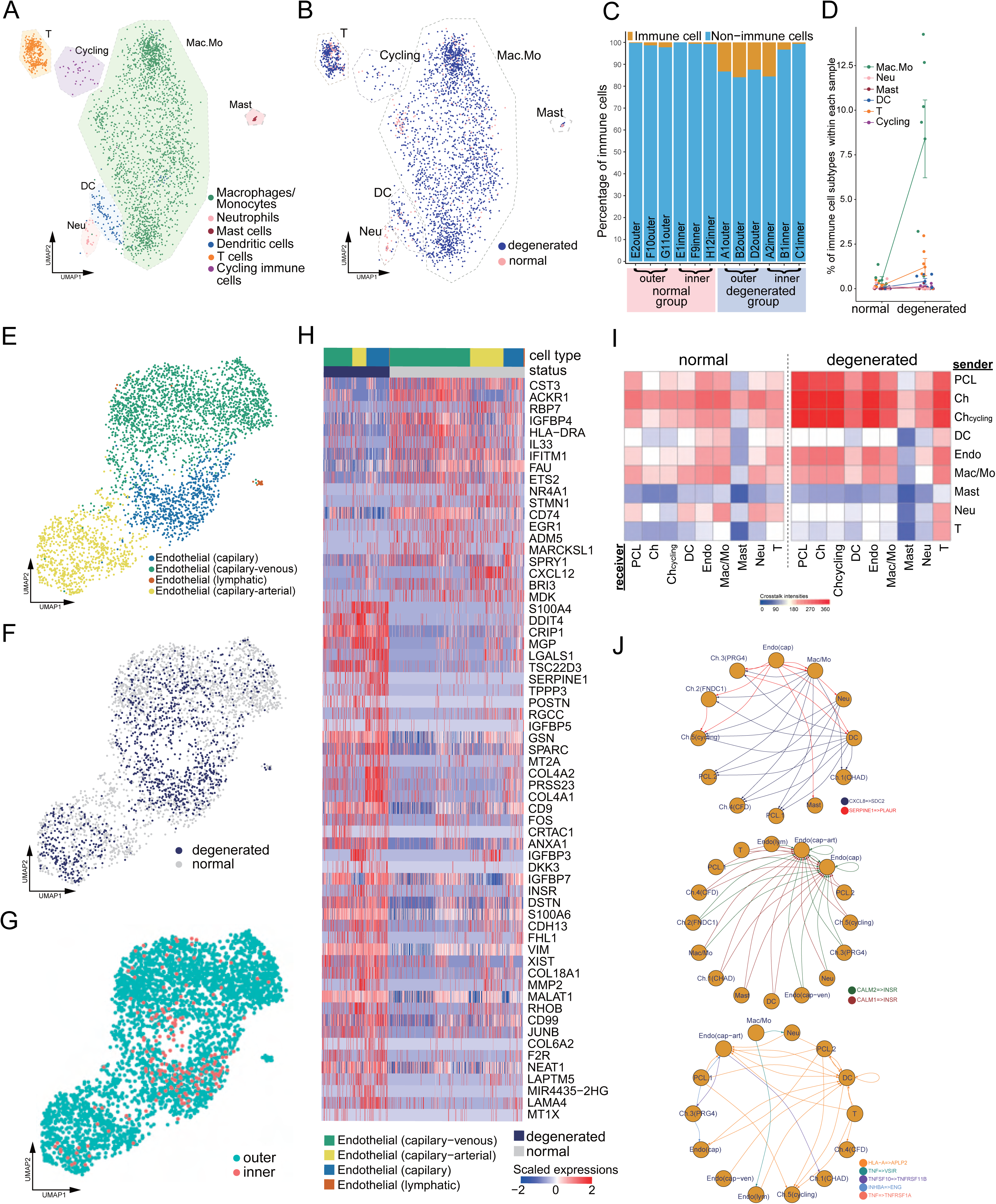
Immune, endothelial cells and their crosstalk with chondrocytes. (A) UMAP visualization of the immune cell-types in the meniscus. (B) UMAP visualization of the statuses of samples (degenerated/normal). (C) Per-sample bar plots visualize the immune cell percentage changes between the normal and degenerated group. (D) Summarized immune cell percentage changes. Error bars show the standard deviations of the data. (E) UMAP visualization of the endothelial class cells’ subtypes. (F) UMAP visualization of sample status. (G) UMAP visualization of the anatomical regions. (H) Top differentially expressed genes between different health states (degenerated vs. normal). The heatmap shows z-score-scaled gene expression values. (I) General cell-cell crosstalk between large populations. The ligand-receptor pair crosstalk was evaluated at the large population level (Methods) (J) Representative crosstalk significantly enhanced in the degeneration group.

Macrophage/monocyte makes up the largest portion of the total immune population. The percentages of macrophage/monocyte, T cell, dendritic cell, and the total immune cell, increased in the degeneration group, which was consistent with the vascularization status and the damage-induced inflammation (Figure 5 B-C). We noticed some proinflammatory cytokine genes and chemokine genes are regulated in the degeneration group, e.g., IL1A, IL1B, IL6, IL15, IL18 in cycling immune cells; CXCL1, CCL3, CXCL3 in macrophages/monocytes (Supplementary Figure S14D). We also noticed increased expressions of metallothionein genes in the degenerated group, e.g., MT1X, MT1G in macrophages/monocytes (Supplementary Figure S14 B), MT1X, MT2A, MT1E in T cells (Supplementary Figure S14C).

We identified an arterial-capillary-venous gradient in the meniscal endothelial cell population (Figure 5E-G). The capillary-arterial endothelial cells highly express GJA5; the capillary endothelial cells highly expressed PLVAP, and the capillary-venous endothelial cells highly expressed VWF. These endothelial cells comprise the microvascular and capillary vessels. A tiny cluster of lymphatic endothelial cells was also observed, which may come from the lymphatic vessels in the meniscus. We compared the total endothelial cells between the normal/degenerated group and reported a list of DEGs (Figure 5H). In the normal status, CST3, ACKR1, RBP7, IGFBP4 were upregulated, while in the degenerated group, S100A4, DDIT4, CRIP1, MGP were upregulated. CST3 is ranked first in the normal group DEGs, which was reported to have inhibition effects on the proliferation, migration, tube formation, and permeability of endothelial cells^36^. ACKR1 is also highly expressed in the normal group, which encodes a chemokine decoy receptor inhibiting the effectiveness of other chemokines. These proteins contributed to the blood vessel’s stability in the normal group.

With all major classes of meniscal cells being identified, we analyzed the cell-cell interactions in the normal/degenerated status and found the crosstalk intensities among cells generally increased in the degeneration group, especially for endothelial cells and immune cells (Figure 5I). These enhanced cell-cell interactions also explained the angiogenesis and leukocyte recruitment phenomena in the degeneration group (Figure 6). In Figure 5J, we illustrated some interactions mediated by ligand-receptor pairs originating from the sender cell type to the receiver type. The intensities of the reported interactions are higher than the background level (Methods). Specifically, in the degenerated meniscus, CXCL8 was produced by macrophages/monocytes, neutrophils, and DC; and its receptor SDC2 was expressed on the chondrocytes and the pericyte like cells. Many cell types produced CALM1/2 that binds to the endothelial cells and promotes the growth of the endothelium^37^. We also identified degeneration group enhanced crosstalk mediated by: class I HLA → APLP2 interactions, TNF superfamily interactions (TNF, TNFSF10 → VSIR, TNFRSF1A/B), and TGF superfamily interactions (INHBA→ENG). In Supplementary Figure S13A- S13L, we illustrated the crosstalk that is enhanced in the degeneration or normal group with detailed interaction sender/receiver cell types and specific ligand-receptor pairs.

**Figure 6:**
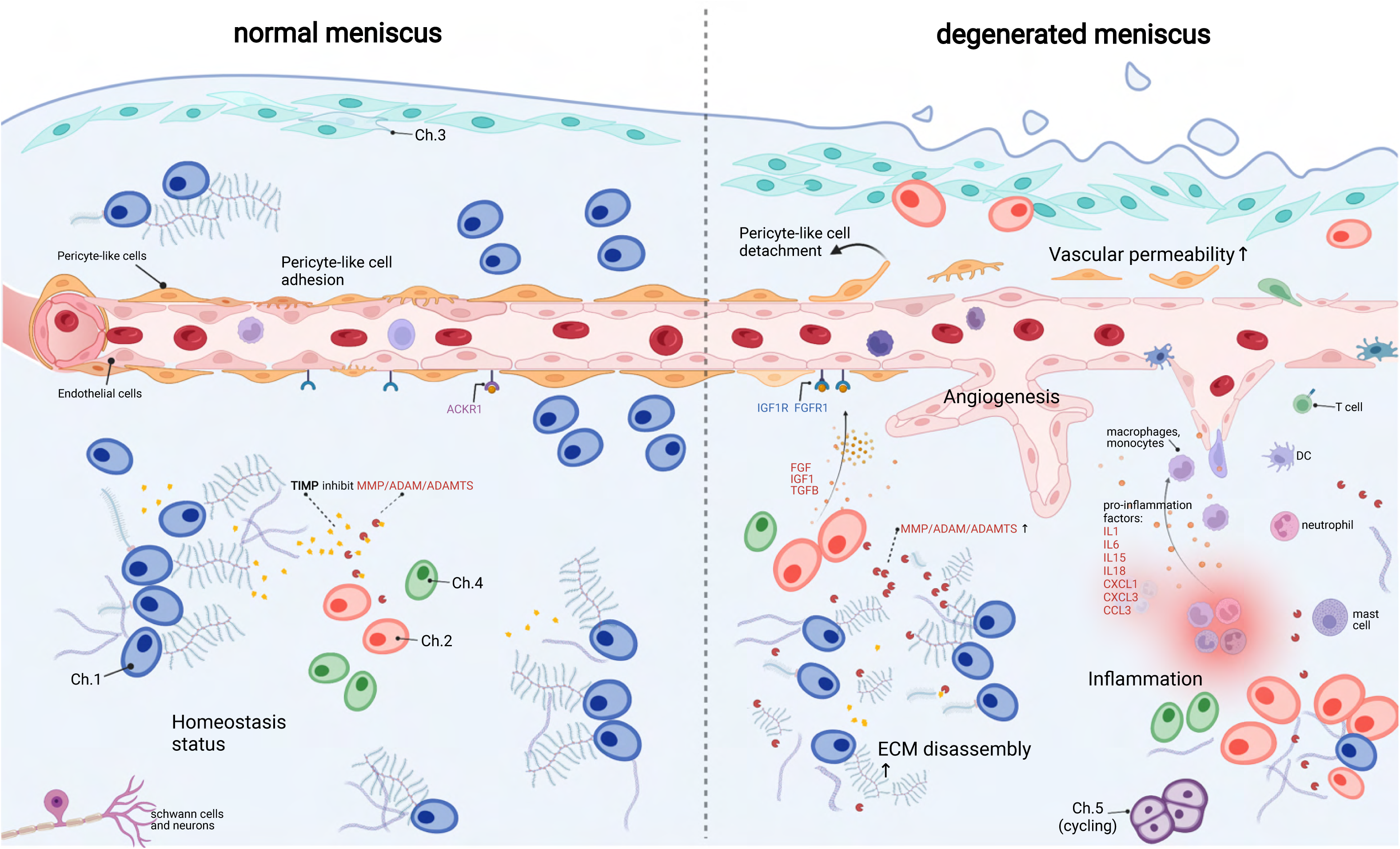
A schematic diagram of the microenvironment changes between the normal and the degenerated meniscus. The left side visualizes the homeostasis meniscus, where Ch.1 is the dominant chondrocyte population. In this situation, the ECM decomposition and synthesis reach a dynamic equilibrium, and the aberrant proliferation of blood vessels is inhibited. The right side visualizes the degenerated meniscus where the orchestrated microenvironment balance is broken. The pericyte-like cells detach from the blood vessels and lower the stability of the blood vessels. The endothelial cells grew and formed new blood vessels in the degenerated area. The vascular permeability increased and allowed more immune cells to infiltrate the degenerated tissue. Ch.2 and Ch.3 increased in quantity and produced more matrix disassembly enzymes, damaging the ECM, releasing angiogenesis factors, and recruiting more immune cells. The immune cells, such as macrophage/monocytes, DCs, T cells, also have chemotaxis functions. There are also cycling chondrocytes and nerve tissue cells in the meniscus.

Based on the above comparisons, we can draw a conclusion that the immune cells and endothelial cells are important regulators for chondrocyte and ECMs (Figure 6). Damages in meniscal tissues broke the ECM and released damage-associated molecular patterns (DAMP) that initiated inflammatory responses. Sensing the DAMP signals, immune cells were then recruited to the inflammatory meniscus and produced cytokines and chemokines, which in turn upregulate catabolic enzymes like MMP/ADAM/ADAMTS and promoted ECM disassembly. The inflammation also stimulated the endothelial cell proliferation and the sprouting new blood vessels further promoted the immune infiltration in the meniscal tissue. All these factors formed a positive feedback loop and drove the microenvironment far away from the homeostasis.

## 3. Discussion

Recent progress in single-cell studies has addressed the importance of meniscal progenitors in tissue engineering ^22^ and gave a rough picture of the meniscal cell types. However, the accumulated data were insufficient to reveal degeneration participants in the microenvironment. In this work, we investigated meniscal chondrocytes, proposed a hierarchical classification system, and classified them into five populations (Ch.1 - Ch.5). We identified endothelial cells enriched in both outer meniscal parts and inner parts, which are typically conceived as avascular “white-zone”. We inferred these unanticipated cells were the degeneration product because newly generated vascular endothelial cells usually occur in cartilage as the osteoarthritis progresses^38^. Surrounding the microvessels are the pericyte-like cells possessing muscle contraction features.

We also observed Schwann cells, which typically wrap around the neurons, indicating there are nerve tissues in the human meniscus. For the immune cells, we observed monocytes/macrophages, DCs, T cells, neutrophils, Mast cells, and some cycling immune cells.

The meniscal microenvironment changes significantly from the homeostasis when the degeneration progresses (Figure 6). In normal meniscal tissues, the synthesis and disassembly of the matrix molecules were balanced in the orchestrated ECM metabolism. We inferred that the dominant Ch.1 population was essential to the balance maintenance because they produced TIMP family molecules that inhibited the ECM decomposers (MMP/ADAM/ADAMTS). The PCL population that adhered to the surface of endothelial cells maintained vasculature stability. The high-level expressions of atypical chemokine receptors like ACKR1 neutralized the chemokines and attenuated the angiogenesis^39^. The patrolling immune cells mainly pass by the tissue in the blood vessel without residing or being attracted.

However, the balanced microenvironment altered significantly in the degenerated meniscal tissues. The secreted ECM decomposers (MMP/ADAM/ADAMTS) from chondral subpopulations Ch.2 and Ch.3 might surpass the inhibition forces (TIMP) and decompose the ECM by breaking large proteoglycan molecules into fragments. We believe the blood vessels have two sides in the homeostasis of the meniscus. On the one hand, the blood vessels help to restore acute injuries to health; on the other hand, the proliferated blood vessels can build pathways for the immune cells so that they can infiltrate the meniscal matrix and cause inflammation. We observed multiple synergistic factors promoting angiogenesis and increased endothelial percentages in the degenerated samples. While the pericyte stabilizes the blood vessel in health, the detachment and decrease of the pericyte may lead to pathological angiogenesis.

On the vascular wall, the endothelial cells downregulated ACKR1, sensed the angiogenesis factors like VEGFA, FGF, IGF1, TGFβ produced by the Ch.2, Ch.3 Ch.4, grew new branches, increased the vascular permeability, and finally allowed more immune cells to enter the matrix and causing inflammation. If the inflammation response, the vascularization process, and the ECM decomposition reached a mutually reinforcing state, the meniscus degeneration would be further aggravated. Changing the chondrocyte subtype ratios and limiting blood vessel proliferation may delay or reverse the meniscus degeneration. In addition to these scientific findings from the analysis of the meniscal single-cell sequencing data, we have also made an interactive website to store the processed data. Readers can directly access the website through the link, and perform some online analysis of the results, comparison of differential genes between two special clusters, data download, etc., which brings great convenience.

During the design of this study, we only partitioned samples into two health states: normal and degenerated. If we distinguished disease severity of the meniscus samples, we could establish more fine-grained links between the molecular profiles and the disease. We observed a small number of nerve tissue cells, which also hindered detailed analysis on them. Our functional coordination of chondrocyte subtypes was more or less confined to the meniscus environment even though there were signs that chondrocytes across different cartilage tissues were not distinct and have many common features. We hope a future molecular coordination framework could integrate chondrocytes across multiple cartilage tissues, different health states, anatomical regions, differentiation stages, guiding the precise taxonomy of chondrocytes when more single-cell cartilage studies were released.

In conclusion, we systematically profiled the cellular diversities in the inner and outer meniscus and reported the microenvironmental alterations in the healthy and degenerated states. Our study provides a great complement to the meniscal single-cell sequencing data and provides an important reference for the study of meniscal degeneration. We should give attention to the blood vessel’s functional duality in acute and chronic damage. Preserving the pericyte-like cells wrapping around the vessels might be a potential strategy to combat the chronic degeneration. The positive feedback loop of the inflammation response, angiogenesis process, and ECM catabolism explained the difficulty in homeostasis restoration. Tunning cells participating in these processes to homeostasis and containing the positive feedback is crucial for delaying or reversing meniscus degeneration. Meniscus is an important and representative fibrocartilage in the human body. We speculated that the cellular and molecular mechanisms observed in this study during meniscal degeneration such as angiogenesis and inflammation could also apply to intervertebral discs and articular cartilage degeneration. It could shed new lights on the diagnosis and treatment of other degeneration-associated musculoskeletal diseases like low back pain and joint pain.

## 4. Methods

### 4.1. Human meniscus cell sample preparation

The meniscus specimens were collected from four patients with meniscus degeneration and four amputation patients with the normal meniscus. The experiments were approved by our University Ethics Committee (Ethics Committee on Biomedical Research, West China Hospital of Sichuan University No. 2020-(921)). The meniscus tissues were removed from the patient’s knee joints and divided into the inner (white zone) and outer (red zone) areas. The degeneration group is comprised of patients A, B, C, D; and the normal group is comprised of patients E, F, G, H. Patient A, B, E, F contributed paired inner-outer samples. Patients C and H contributed unpaired inner samples, and patients D and G contributed unpaired outer samples because the other pair had obtained in the surgery had poor sample states. Detailed designs of experiments can be found in Supplementary Table S1.

After carefully dividing the meniscus tissue into the inner and outer parts, the menisci were cut into small pieces. We used 0.25% Trypsin-EDTA (Gibco 25200072) to digest the pieces for 1 hour under 37℃. Next, we used 4 mg/ml collagenase IV (Sangon Biotech A004210) to dissociate the pieces for 6 hours under 37℃ and finally obtained dissociated cells. The dissociated cells were resuspended at a concentration of 250-1200 cells/μL and with viability between 67%-87% for microfluidics (Chromium Single-Cell Controller, 10x Genomics). According to the manufacturer’s instructions, cells were loaded into the chip and run using the Chromium Single Cell 3′ Reagent Kit v2 (10x Genomics). Sequencing-ready single-cell transcriptome libraries were mapped to the human reference genome GRCh38-3.0.0 and quantified by CellRanger 3.1.0.

### 4.2. Multiplex immunofluorescence (OPAL™) staining

Human meniscus normal and degenerated samples were fixed in 35% paraformaldehyde and embedded in paraffin. We sliced the embedded paraffin samples in series, and each slice was 4 µm thick. Firstly, water-bath heating was used for antigen retrieval. Then various cell marker primary antibodies were incubated with the paraffin slide of meniscus tissue to conduct the continuous staining with the Opal Polaris Multiple-Color Manual IHC Kit (NEL861001KT). We used different primary antibodies to simultaneously label Ch.1(SERPINA1), Ch.2(MMP14), Ch.4 (CYP1B1), PCL(ACTA2) on the same tissue slide. The automated staining system (BOND-RX, Leica Microsystems, Vista, CA) was used to perform the chromogen-based multiplex immunohistochemistry labeling. SERPINA1 (ab207303), MMP14 (ab51074), CYP1B1 (ab33586), and ACTA2 (ab5694) were all purchased from Abcam and diluted at a concentration of 1:100. The Opal Polaris dyes were used to pair with these antibodies containing fluorophores for tyramide signal amplification (TSA) to enhance sensitivity. The sequence of labeling for detecting each marker was optimized: CYP1B1 (Opal 570), MMP14 (Opal 620), SERPINA1 (Opal 690), ACTA2 (Opal 780), and DAPI. Including an autofluorescence section, the staining process is the same as above, but no primary antibody is added. Multiplex analysis was operated to analyze the results of simultaneously stained tissue slides. In addition, common immunofluorescence staining was performed using CDON (ab227056), CD31 (ab9498), and CD45 (ab40763) labeled Ch.3, endothelial cells, and immune cells. All experiments were biological replicated 3 times in the laboratory.

### 4.3. Single-cell data quality control steps

We adjusted the ambient RNA expression with SoupX ^40^ v1.2.1 and generated read counts free of background noises for all downstream analyses. We used scanpy^41^ and Seurat^42, 43^ to conduct the basic filtering out outliers cells with high n_genes metrics (the number of observed genes per cell), high n_counts metrics (the number of UMI per cell), and high percent_mito metrics (the mitochondrial gene fraction). Next, we assigned draft cell type identities to the cells using SingleR^44^. Note that the SingleR-generated cell type labels were only used to perform quality control analysis to avoid removing biologically meaningful cells with poor data qualities. We calculated doublet scores using DoubletFinder^45^ removed the cells with high doublet scores and outlier clusters with high doublet scores. We observed that DoubletFinder tends to identify small clusters as doublets incorrectly, so we assert these small populations as singlets if they had well-defined identities given by SingleR or appear clear biological functions such as cell cycles. To remove the confounding factor introduced by the dissociation-induced gene expression^46, 47^, we decided to remove the heavily affected cells by the dissociation steps. We calculated dissociation scores using methods described in van den Brink, et al. ^46^.

Raw single-cell RNA-seq datasets contain numerous low-quality droplets. Hence, we should ensure the majority of the data items we analyzed correspond to viable cells. It is known that one simple quality control filtering on the aforementioned metrics (n_genes, n_counts, percent_mt) can’t remove all of them. We then conducted comprehensive quality control steps that can be summarized as 1) top-down refinements 2) divide and conquer. First, we divide cells into several rough populations with distinct biological identities, for example, chondrocytes, immune cells, endothelial cells, etc. We zoomed into these populations one by one and performed sub-clustering within them. The within-population sub-clustering often works like a centrifuge – the low-quality cells mixed in the large rough population usually form tiny outlier groups when sub-clustering is performed on a single population. When a tiny group is mainly comprised of cells with high doublet scores, cells with high mitochondrial gene fractions, cells with high dissociation scores, we consider removing it. After manually inspecting and dropping these tiny outlier groups, we finally get much cleaner datasets.

### 4.4. Single-cell data clustering steps

To eliminate the genetic background variations and technical noises of the single-cell RNA-seq data, we used harmony ^48^ to perform integrative clustering across different samples. Major clusters corresponded to chondral cells, ACTA2+ cells, endothelial cells, immune cells were identified across samples and validated with cluster correlation analysis (Figure S1C and S4A).

To understand large cell populations with higher resolutions, we subset the chondral and ACTA2+ majors clusters, performed re-clustering on harmony derived embeddings, and obtained multiple fine clusters. To alleviate the scRNA-seq data noise and promote data consistency across samples ^49^, we partitioned original cell data points into homogenous *metacells* with MetaCell ^50^.

We then used HGC ^51^ to build hierarchical relationships based on the *metacells* and summarized the results in Figure 2D.

### 4.5. Differentially expressed genes (DEG) analysis

Differentially expressed genes (DEG) were identified by the “FindMarkers” function and the “FindAllMarkers” function provided by the Seurat package.

The DEGs of each chondrocyte cluster were identified by comparing the cluster with all the other clusters. The health status (degeneration/normal) markers were calculated in an overall comparison way and a per-cluster comparison way. We first extracted the total chondrocytes (from Ch.1 to Ch.5) and compared them between the degenerated and normal groups. We made the same comparison to the total PCL populations. The health status markers were reported in Supplementary Figure S6A. We also performed a health status comparison within each cluster).

### 4.6. Gene set enrichment analysis and gene set variation analysis

The enrichment analysis was performed using DEGs with top fold-changes and adjusted p-value <0.05, with the help of the clusterProfiler^25^ package.

Significantly enriched ontology terms of each cluster (Ch.1-5, PCL.1-2) were obtained using top cluster markers. Significantly enriched ontology terms of the healthy and degenerated status were displayed in Supplementary Figures S6B and S6C. The tree visualization of the enriched terms was done with the help of the treeplot function provided by the enrichplot R package^25^.

We assigned gene set activity scores to individual cells using GSVA ^26^, with gene sets obtained from MSigDB ^52, 53^. The GSVA derived gene set scores were converted to z-scores and visualized in Figure 2G and Figure 3C-3F. The chondrocyte cluster gene set variation analysis (radar plots in Figure 2G) were calculated using the following MSigDB terms respectively:

“GOBP_LEUKOCYTE_MIGRATION”, “GOBP_NEGATIVE_REGULATION_OF_VASCULAR_ENDOTHELIAL_CE LL_PROLIFERATION”, “GOBP_POSITIVE_REGULATION_OF_VASCULATURE_DEVELOPMENT “, “GOBP_RESPONSE_TO_BMP”, “GOBP_COLLAGEN_FIBRIL_ORGANIZATION”, “GOBP_RESPONSE_TO_INTERLEUKIN_1”, “GOBP_RESPONSE_TO_TRANSFORMING_GROWTH_FACTOR_BETA”, “GOBP_RESPONSE_TO_TUMOR_NECROSIS_FACTOR”, “GOBP_MYOFIBRIL_ASSEMBLY”.

### 4.7. Gene regulation network (GRN) inference

We used the python package pySCENIC^27, 28^ (GRNboost2) to infer gene regulatory networks from single-cell expressional profiles. The workflow includes three sub-steps: identifying co-expression modules, trimming modules with prior knowledge, and calculating activating scores.

The workflow builds co-expression modules by choosing TFs that predict target gene expression well. First, the algorithm builds gradient-boosting machines models predicting each target gene’s expression from TF expressions and keeps the models’ feature weights as a measure of TF-target regulatory scores. Second, the algorithm creates modules by adding/filtering weighted links between TFs and their targets. To reduce stochasticity, the algorithm independently uses six weight filtering rules to build modules. The six rules are: 1) keep TF-target pairs with weights > the 75% percentile of the weights; 2) keep TF-target pairs with weights > the 90% percentile of the weights; 3) for each TF, keep top 50 TF-target pairs with the highest weights; 4) for each target, keep top 5 TF-target pairs with highest weights; 5) for each target, keep top 10 TF-target pairs with highest weights; 6) for each target, keep top 50 TF-target pairs with highest weights. After this step, many modules that contain a TF and its targets are created. The algorithm only keeps TF-target links that have positive regulation relationships (the Pearson correlation of TF and target expressions > 0.03) and drops modules with <20 genes inside (TF itself included). Finally, we get a list of primary co-expression modules, each containing a TF and its positively regulated targets.

The module trimming process combines the regulatory motif information. We adopt the 10kb up/downstream regions of a gene’s TSS as the gene’s distal enhancer occurrence region and the 500bp upstream/ 100bp downstream region as the gene’s proximal promoter occurrence region. It is more likely that regulatory elements lie in these regions. Hence, with these TSS surrounding settings, we can derive a collection of regulatory regions for a gene set. The cisTarget database provided a list of motifs that are annotated to the corresponding TFs. The database also stored precomputed hidden Markov model-derived motif ranking scores. The algorithm first selects motifs that are enriched in the regulatory regions of a gene set (NES>3.0 and annotated to TF). Next, for each gene set, the algorithm keeps the target genes that are regulated by the selected motif. Finally, the algorithm merges modules with the same TF and constructs TF-target modules called regulons.

To enhance the robustness of the GRN inference results, we repeated this process multiple times and eventually accepted the consensus of the network nodes and links of all runs. The network structure was visualized using the R igraph package^54^. Finally, we quantified and binarized the activation levels of the TF-target modules with AUCell^27^ and visualized them with ComplexHeatmap^55^.

### 4.8. Cell crosstalk analysis

Given the averaged normalized expression value of a ligand gene in a sender cluster *i* as *E*_*L*,*i*_, and that of a receptor gene in receiver cluster *j* as *E*_*R*,*j*_, we defined the crosstalk between cluster *i* and cluster *j* via the ligand-receptor pair L-R is their product *E*_*L*,*i*_*E*_*R*,*j*_. We enumerated all ligand and receptor genes provided by CellTalkDB^56^ and calculated crosstalk products for sender-receiver cluster pairs. We empirically evaluated the statistical significance of this product by counting the more extreme null products after shuffling the cluster labels.

The overall crosstalk level between the sender cluster *i* and the receiver cluster *j* is defined as ∑_*L*_ ∑_*R*_ *E*_*L*,*i*_*E*_*R*,*j*_ if *E*_*L*,*i*_*E*_*R*,*j*_ > *θ*, and *θ* = 1.5 in our cases.

We compared the crosstalk variations between the degenerated and the normal group by subtracting the products in two conditions (visualized in Supplementary Figure S13). We identified ligand-receptor interactions that were high in the normal/degenerated group and grouped the differential interactions into ECM, TNF, TGFb, chemokine, cytokine antigen-presentation categories (Supplementary Figures S13 A-L).

### 4.9. Online cell browser construction

We used the python package cellxgene to provide online cell browser service. The matrices along with other metadata were saved in scanpy h5ad format for cellxgene.

## Data Availability

Data are available in a public, open access repository. The single-cell RNA-seq data, cluster annotations are available at GSA for human (https://ngdc.cncb.ac.cn/gsa-human/) with the accession number PRJCA008120

## Author Contributions

WF, SC, JL, and XZ conceived the project. WF and SC designed the experiments. SC and HG performed bioinformatic and statistical analyses. RY performed the cell and molecular experiments. SC and RY wrote the manuscripts. All authors commented and revised the manuscripts.

## Acknowledgements

The authors are grateful to Dr. Tao Wu from the Institute of Immunology, Tsinghua University, for his helpful discussions. This study was funded by the National Natural Science Foundation of China (81972123, 82172508, 62050178, 61721003) and the National Key Research and Development Program of China (2021YFF1200901).

---

**Supplementary figure S1:**
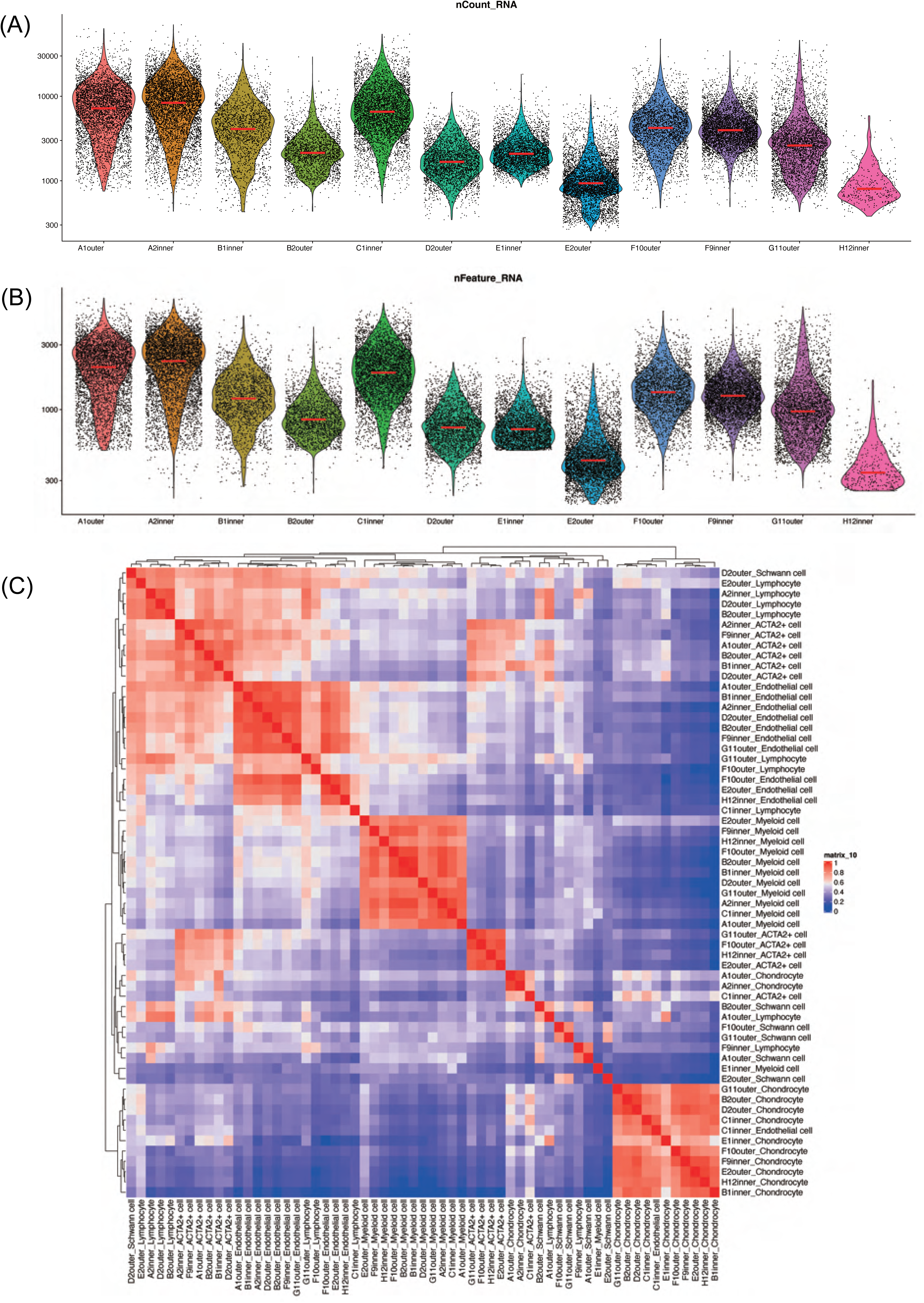
Quality control metrics for single-cell RNA sequencing. (A) Jittered violin plots show the number of UMI counts observed per cell, grouped by the sample. (B) Jittered violin plots show the number of features observed per cell, grouped by the sample. (A) and (B) show that most samples have good sequencing qualities. (C) Cluster expression Pearson correlations. Similar cell types were mostly grouped together, indicating that the cell identities were properly assigned and the batch effects were not significant.

**Supplementary figure S2:**
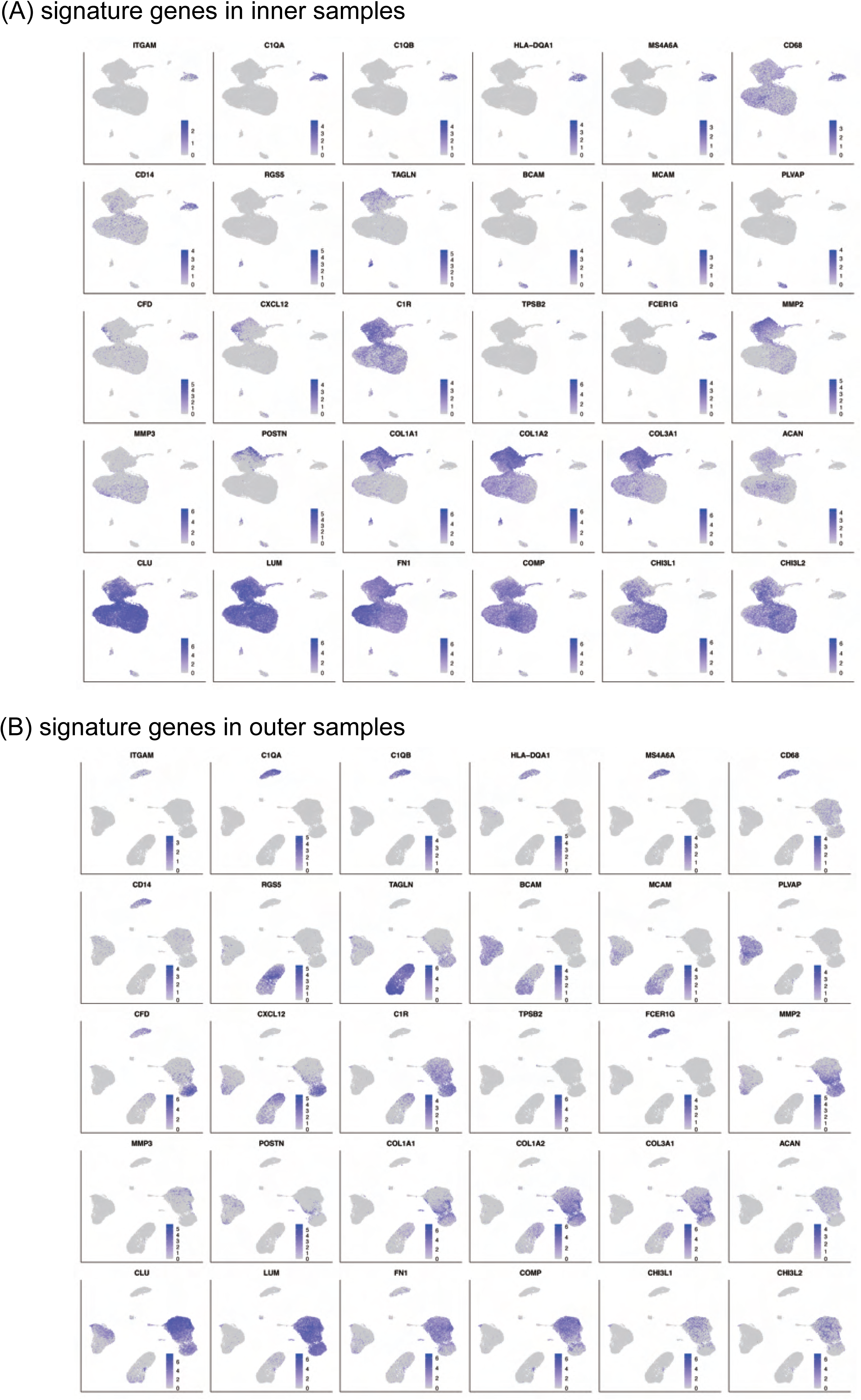
Signature genes of the large cell classes. (A) and (B): The cluster marker genes expression levels are shown on the UMAP plots with colors. Inner samples: up; outer samples: down.

**Supplementary figure S3:**
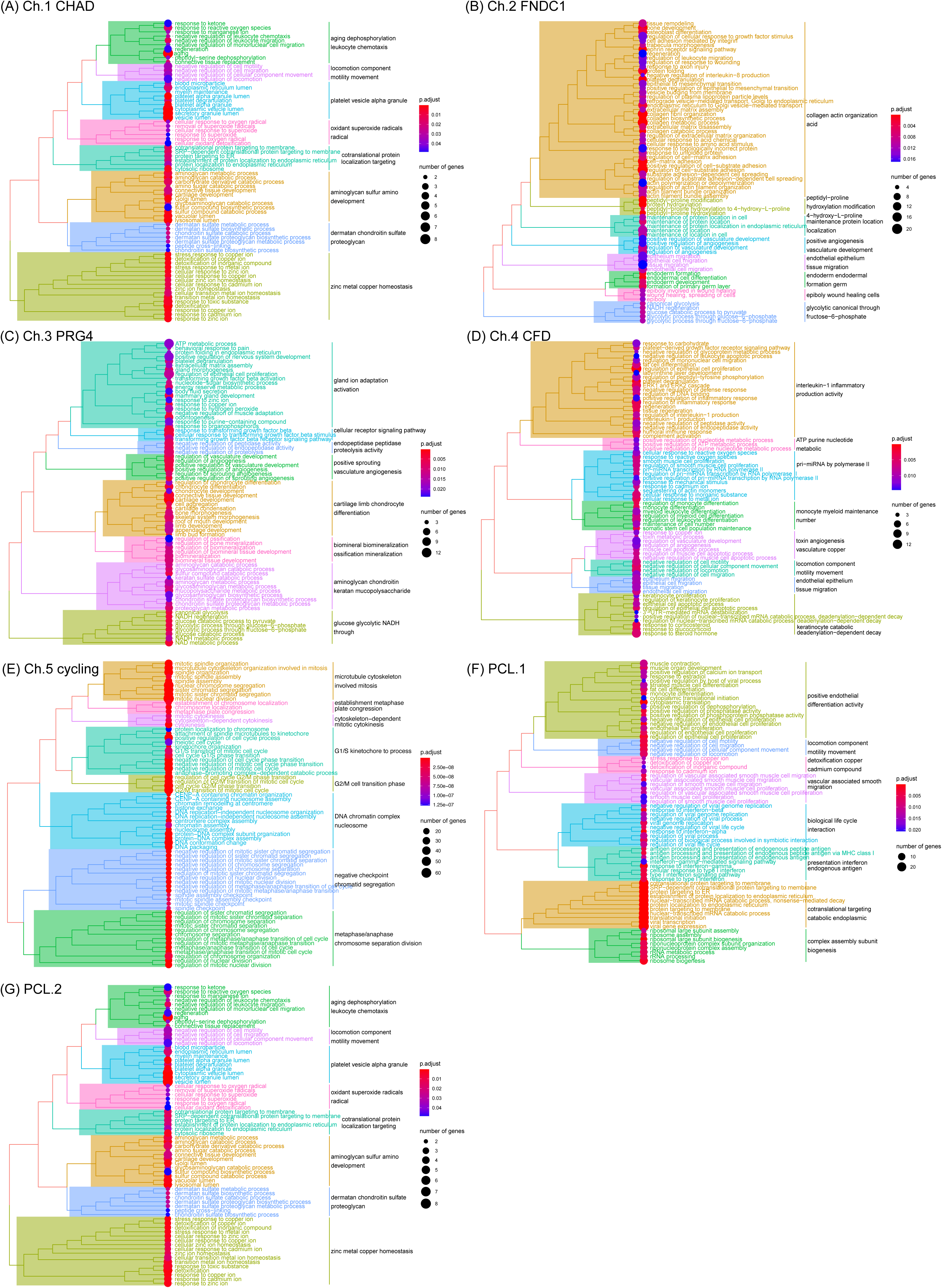
GO enrichment of chondrocyte and PCL subcluster markers. (A)-(G): Pathway enrichment analysis of 7 subpopulations of chondrocytes and pericyte-like cells. The enrichment was performed using gene set over representation analysis using clusterProfiler.

**Supplementary figure S4:**
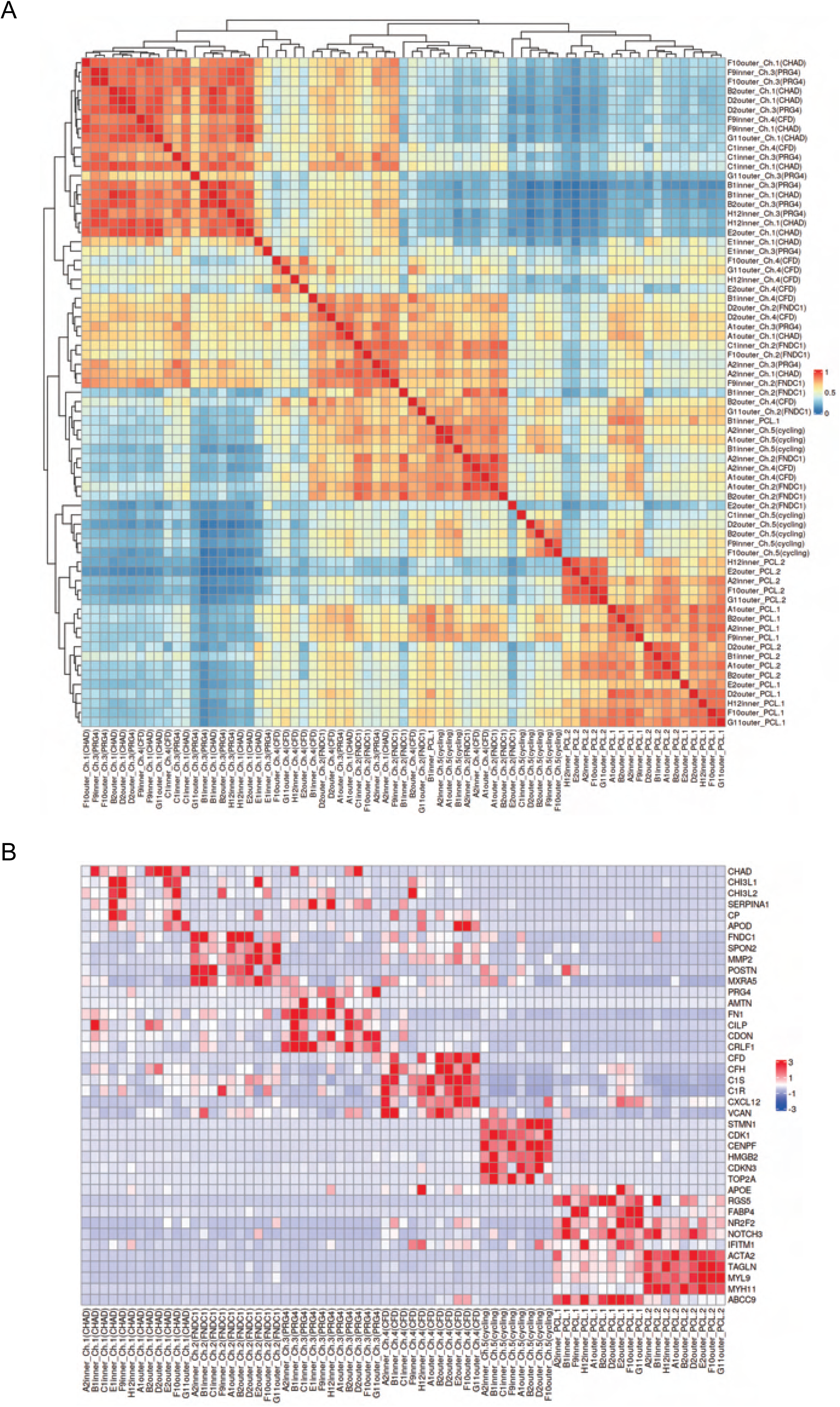
Label agreements of the chondrocyte and pericyte-like cells across samples. (A) Pearson correlations of chondrocyte/PCL subclusters’ expression across all meniscus specimens. (B) The expressions of subcluster signatures genes of across all meniscus specimens. The two heatmaps shows clusters with the same name across different specimens are correlated and share similar expressions.

**Supplementary figure S5:**
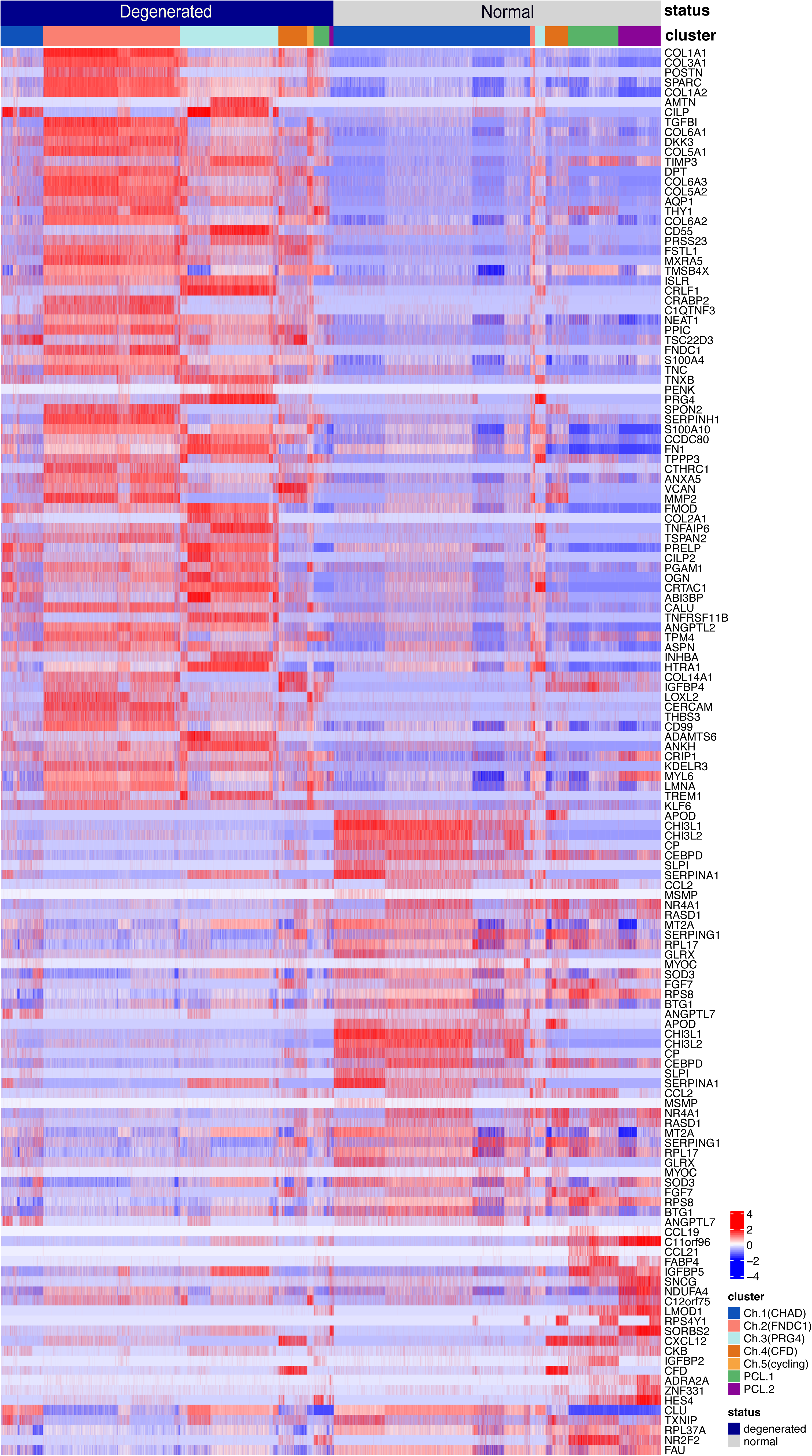
Finding DEGs in total chondrocytes and PCLs (degenerated vs. normal) Heatmap shows the z-score scaled expressions of differentially expressed genes (DEGs) in the degeneration group and the normal group. The DEG analysis was conducted on the total chondrocytes and total pericyte-like cells. Cell types and health statuses were annotated to the top of the heatmap.

**Supplementary figure S6:**
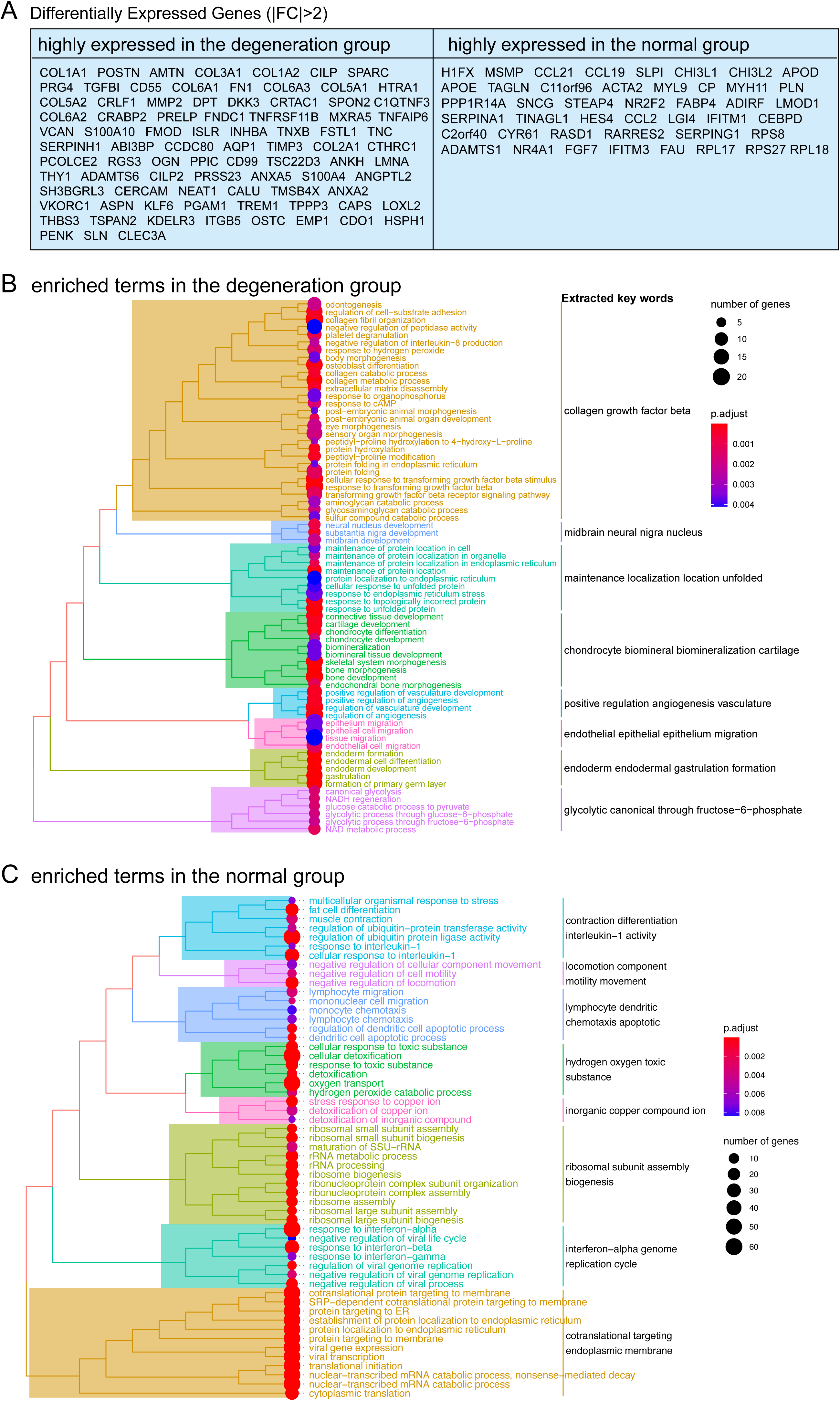
Interpreting DEGs in total chondrocytes and PCLs (degenerated vs. normal) (A) Lists of selected top DEGs with absolute fold changes >2. (B) Enrichment analysis of DEGs in (A). The enrichment was performed using gene set over- representation analysis using clusterProfiler.

**Supplementary figure S7:**
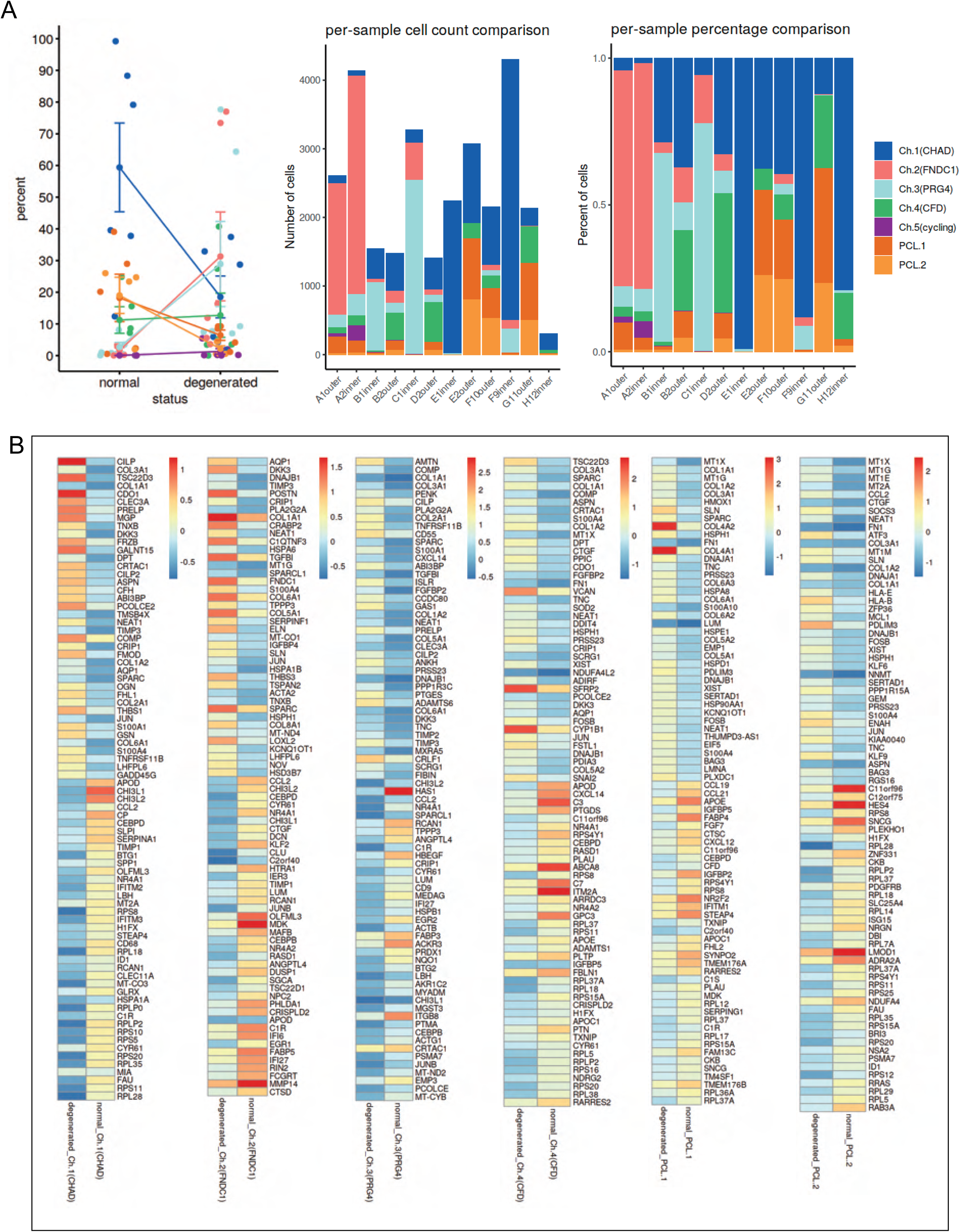
Finding composition changes and DEGs in individual chondrocyte and PCL clusters (degenerated vs. normal) (A) Changes in the composition of the seven chondrocyte subclusters of a meniscus in different statuses and different samples. Left: summarized changes, error bars stand for standard deviations. Middle: per-cluster changes, absolute cell counts. Right: per-cluster changes, relative ratios. (B) Analyses of DEGs between normal and degenerated statuses in an individual cluster.

**Supplementary figure S8:**
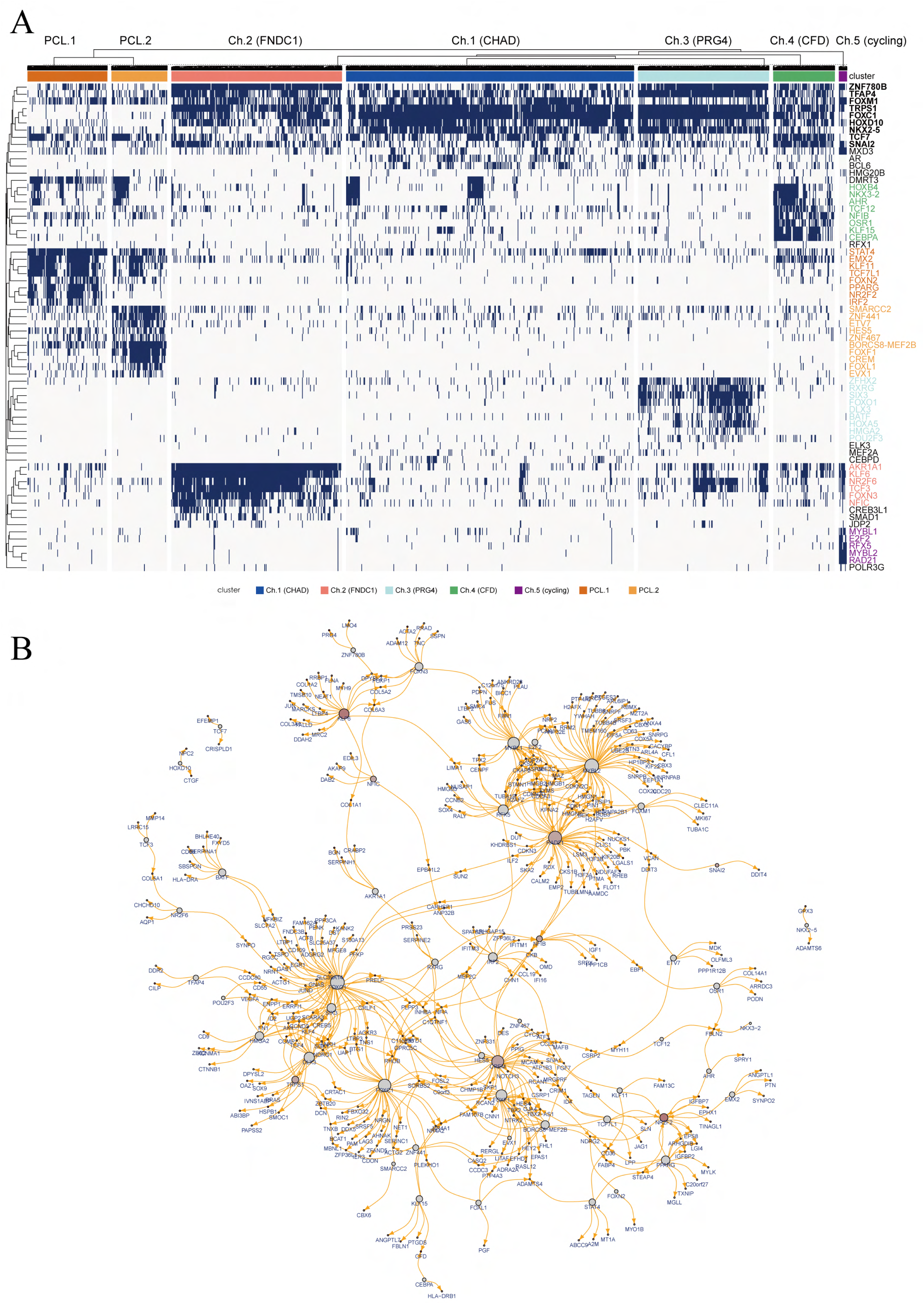
Regulons derived by pySCENIC. (A) Binary regulon activities of chondrocytes and pericyte-like cells. Each row represents a gene regulatory module containing a TF and its targets. The activation status of the module in one cell is converted to ON/OFF states and shown in the heatmap. (B) A core subset of the full gene regulatory network with high expression levels.

**Supplementary figure S9:**
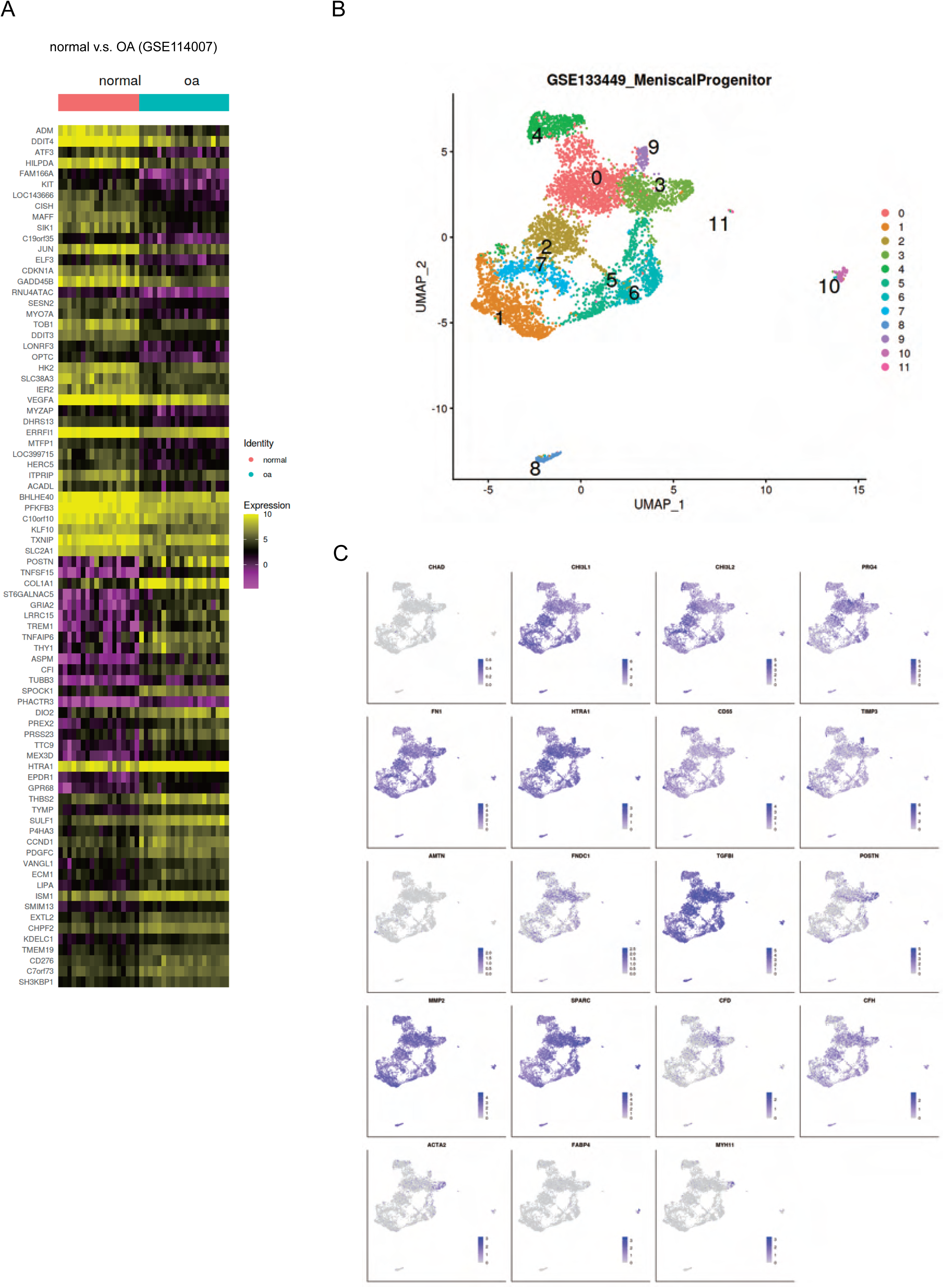
Public chondrocyte datasets reanalysis (I) (**A**) OA-related genes. The heatmap shows the differential gene expression of articular cartilage in normal and OA states by reanalyzing public data (GSE114007). (B) UMAP shows cell clustering results of the scRNA-seq meniscus dataset from Sun et al. (GSE133449). (C) The expression of signature genes in figure 2 on GSE133449. Some highly expressed genes such as CHAD were not detected on their dataset. This might be caused by different single-cell sample preparation protocols: we used the 10x Genomics platform, and they used the BD Rhapsody platform.

**Supplementary figure S10:**
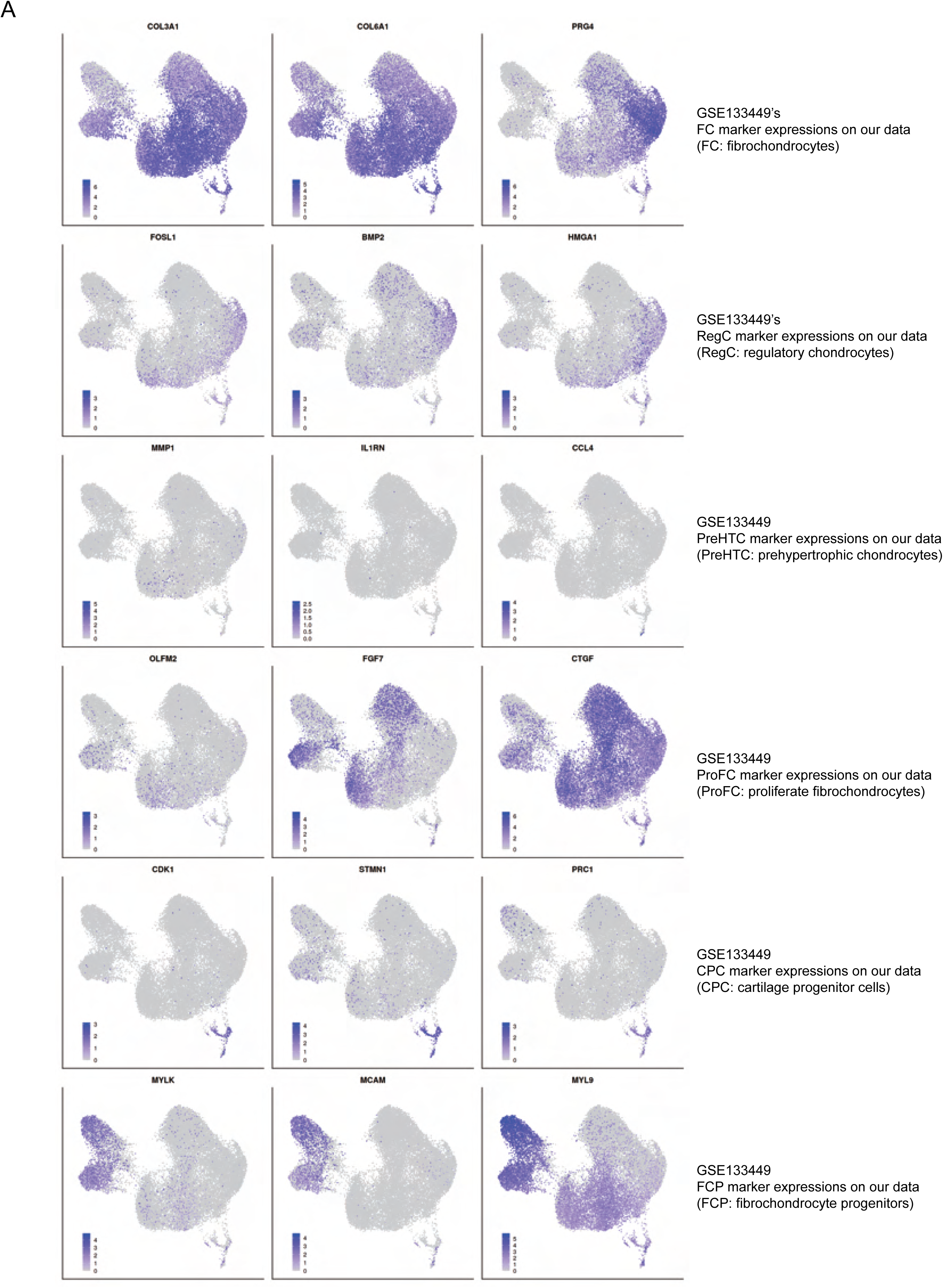
Public chondrocyte datasets reanalysis (II) (A) GSE133449’s normal group marker genes of FC, RegC, PreHTC, ProFC, CPC, and FCP on our data.

**Supplementary figure S11:**
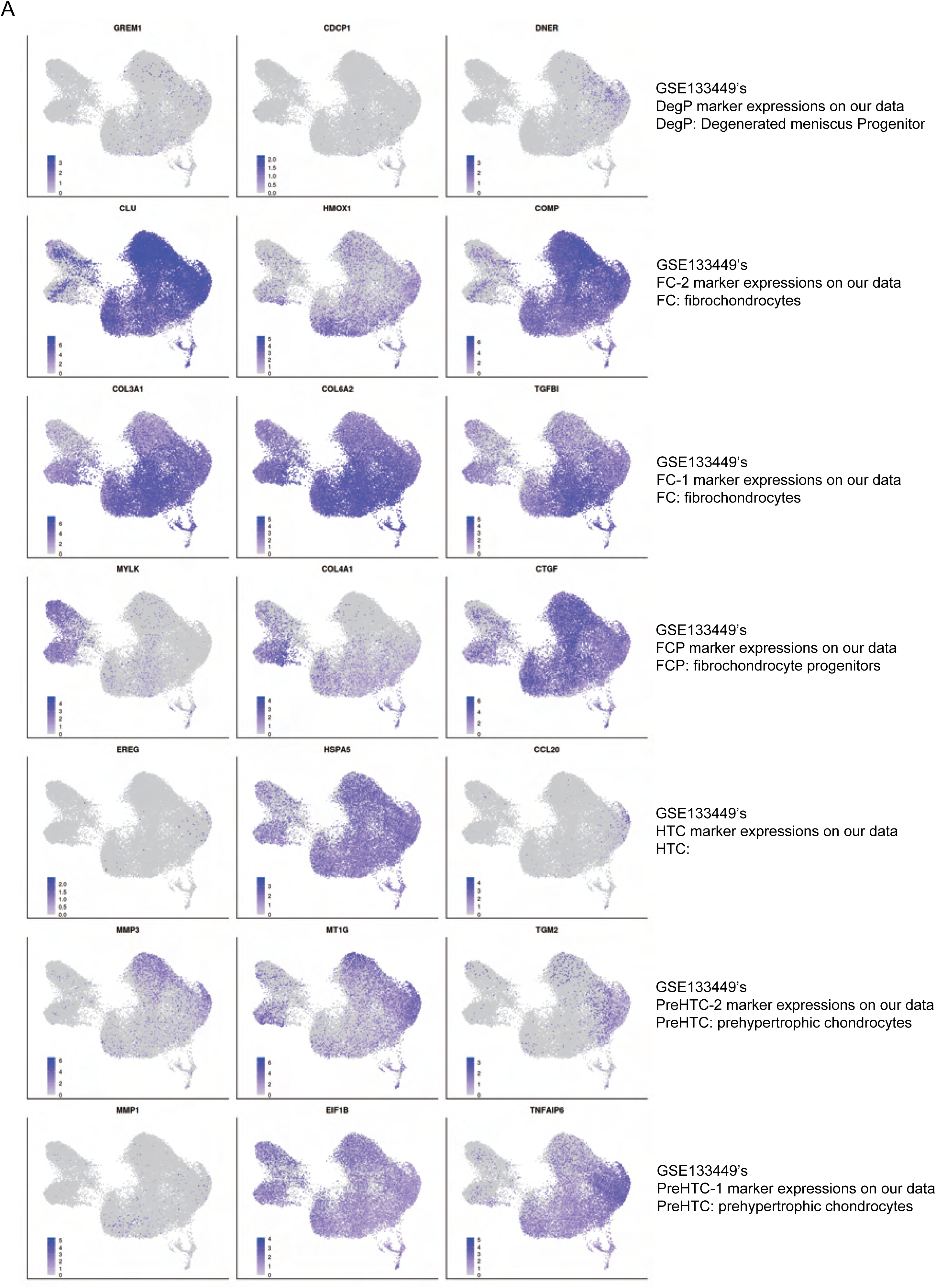
Public chondrocyte datasets reanalysis (III) (A) GSE133449’s degeneration group marker genes of DegP, FC-2, PreHTC, ProFC, CPC, and FCP expressed on our data.

**Supplementary figure S12:**
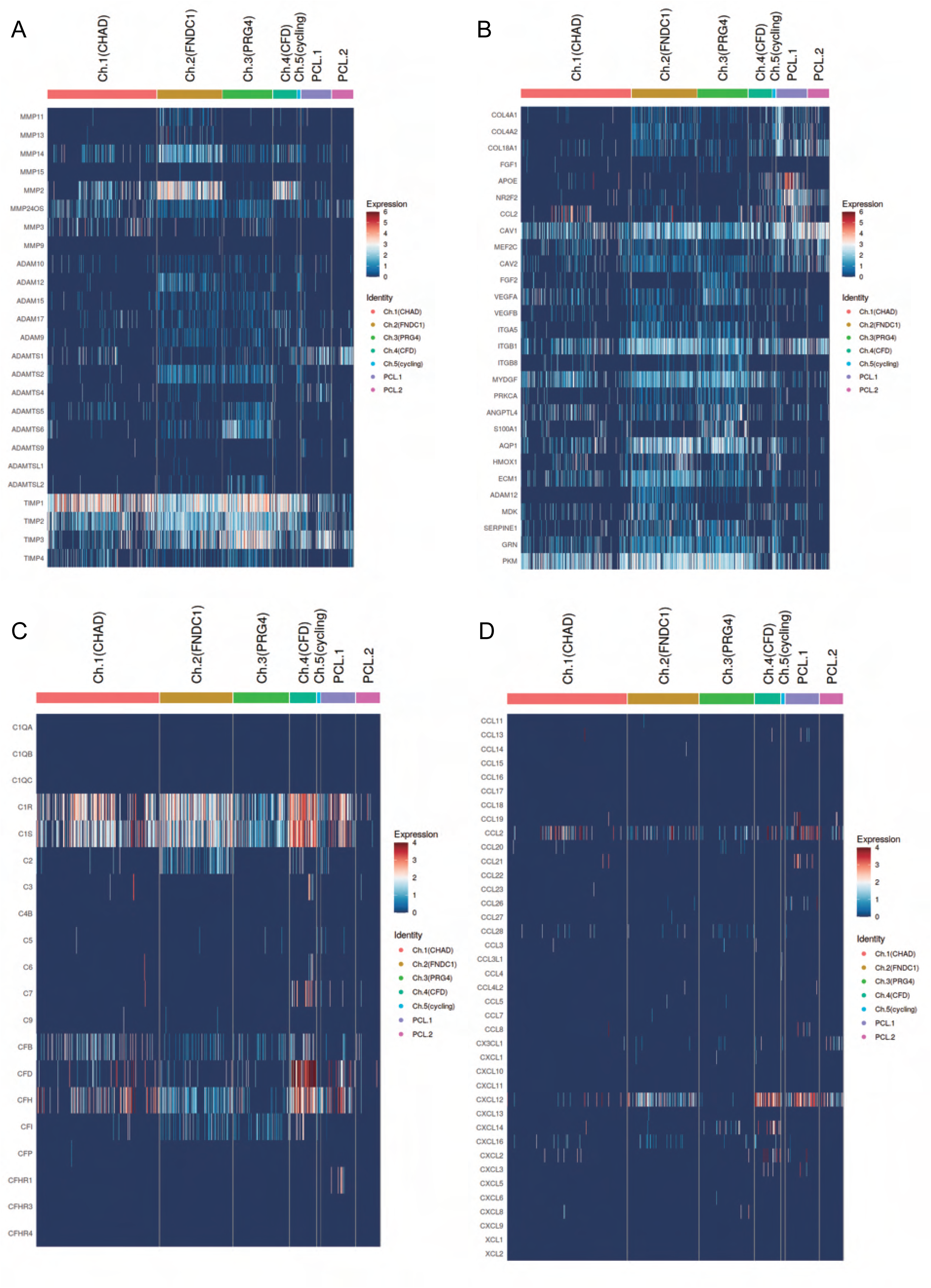
Expression levels of selected gene sets. (A) Expression of ECM decomposition-related genes and anti-decomposition-related genes in chondrocyte subpopulations. (B) Expression of genes associated with angiogenesis and anti-angiogenesis in chondrocytes subpopulations. (C) Expression of complement-related genes in chondrocyte subpopulations. (D) Expression of chemotaxis-related genes in chondrocyte subpopulations.

**Supplementary figure S13:**
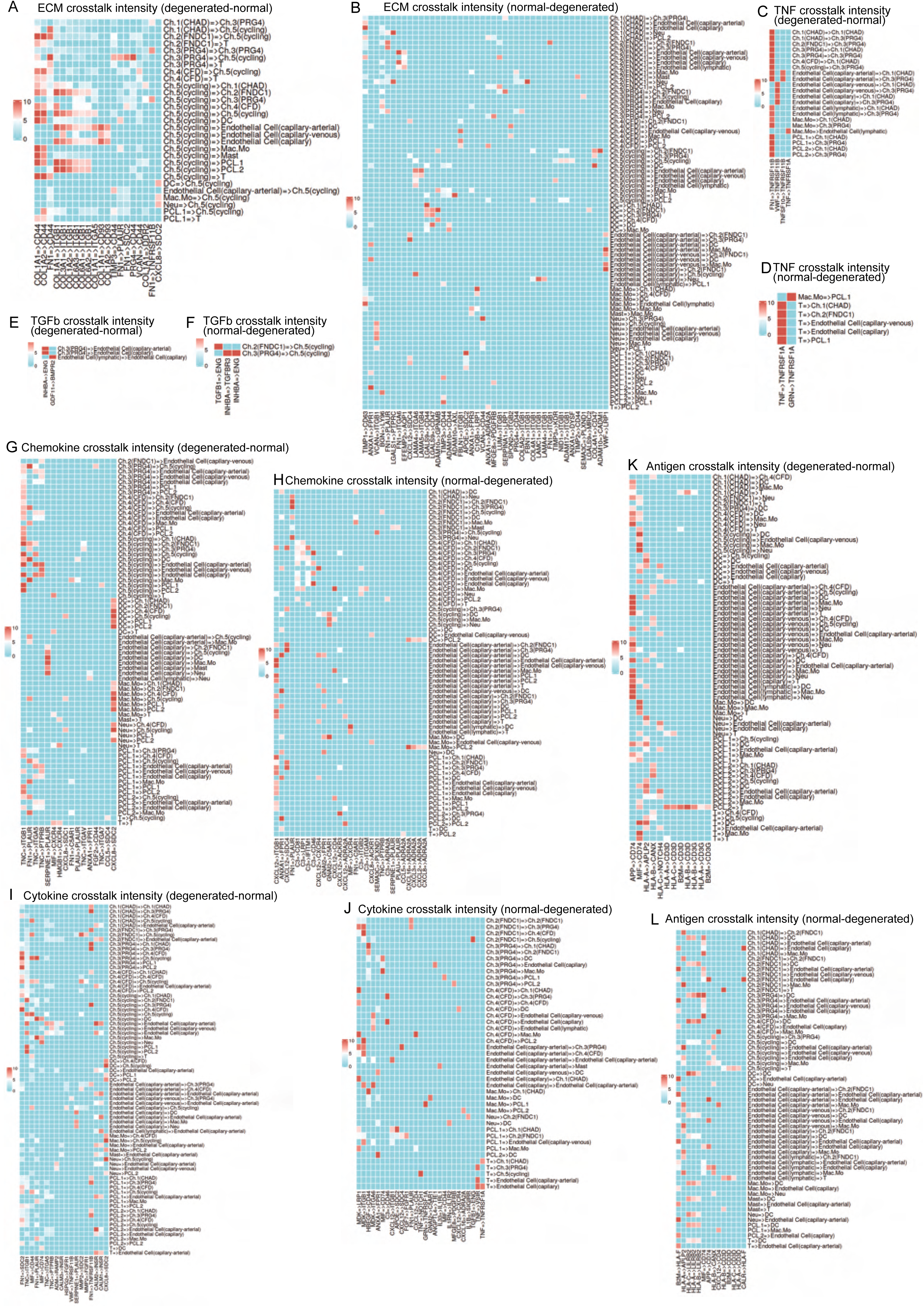
Ligand-receptor pairs that were up/down- regulated in degeneration. **(A)-(L)**: The heatmaps reported ligand-receptor pairs that have significant differences across all sender and receiver cell types

**Supplementary figure S14:**
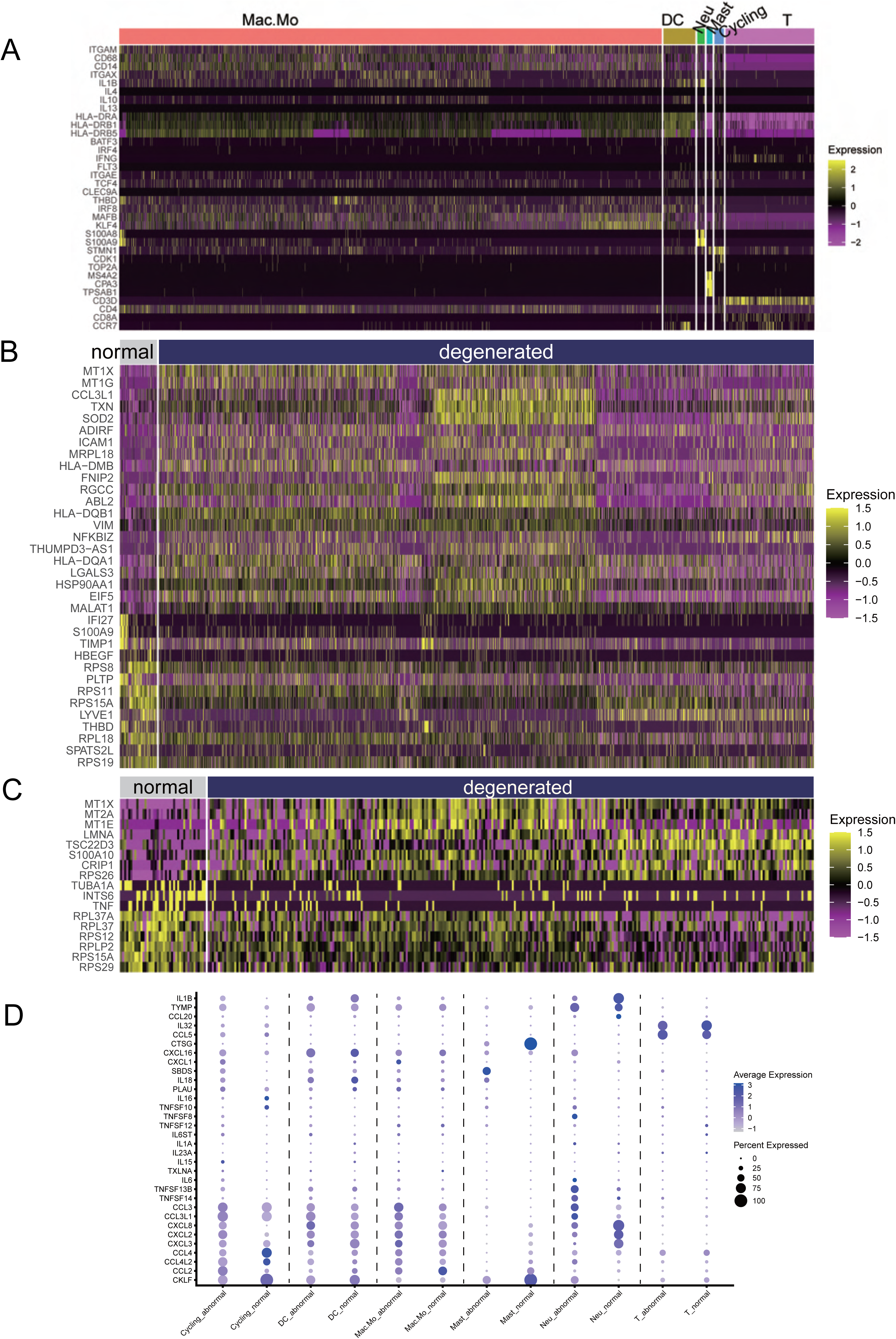
Immune signature genes. (A) Signature genes that were used to define the immune cell subpopulations. (B) Finding DEGs in macrophages/monocytes (normal vs. degenerated). (C) Finding DEGs in T cells (normal vs. degenerated) (D) Analysis of cytokines and chemokines released by various immune cells in the normal and degenerated meniscus.

## References

1. Murphy, C. A. et al. The Meniscus in Normal and Osteoarthritic Tissues: Facing the Structure Property Challenges and Current Treatment Trends. Annual review of biomedical engineering 21, 495–521, doi:10.1146/annurev-bioeng-060418-052547 (2019).

2. Newman, A. P., Anderson, D. R., Daniels, A. U. & Dales, M. C. Mechanics of the healed meniscus in a canine model. The American journal of sports medicine 17, 164–175, doi:10.1177/036354658901700205 (1989).

3. Rai, M. F. & McNulty, A. L. Meniscus beyond mechanics: Using biology to advance our understanding of meniscus injury and treatment. Connective tissue research 58, 221–224, doi:10.1080/03008207.2017.1312921 (2017).

4. Englund, M., Roemer, F. W., Hayashi, D., Crema, M. D. & Guermazi, A. Meniscus pathology, osteoarthritis and the treatment controversy. Nat Rev Rheumatol 8, 412–419, doi:10.1038/nrrheum.2012.69 (2012).

5. Lohmander, L. S., Englund, P. M., Dahl, L. L. & Roos, E. M. The long-term consequence of anterior cruciate ligament and meniscus injuries: osteoarthritis. The American journal of sports medicine 35, 1756–1769, doi:10.1177/0363546507307396 (2007).

6. Sun, Y. et al. Analysis of meniscal degeneration and meniscal gene expression. BMC musculoskeletal disorders 11, 19, doi:10.1186/1471-2474-11-19 (2010).

7. Makris, E. A., Hadidi, P. & Athanasiou, K. A. The knee meniscus: structure-function, pathophysiology, current repair techniques, and prospects for regeneration. Biomaterials 32, 7411–7431, doi:10.1016/j.biomaterials.2011.06.037 (2011).

8. Danzig, L., Resnick, D., Gonsalves, M. & Akeson, W. H. Blood supply to the normal and abnormal menisci of the human knee. Clinical orthopaedics and related research, 271–276 (1983).

9. Herwig, J., Egner, E. & Buddecke, E. Chemical changes of human knee joint menisci in various stages of degeneration. Annals of the rheumatic diseases 43, 635–640, doi:10.1136/ard.43.4.635 (1984).

10. Cheung, H. S. Distribution of type I, II, III and V in the pepsin solubilized collagens in bovine menisci. Connect Tissue Res 16, 343–356, doi:10.3109/03008208709005619 (1987).

11. Yanagishita, M. Function of proteoglycans in the extracellular matrix. Acta Pathol Jpn 43, 283–293, doi:10.1111/j.1440-1827.1993.tb02569.x (1993).

12. Nakano, T., Dodd, C. M. & Scott, P. G. Glycosaminoglycans and proteoglycans from different zones of the porcine knee meniscus. J Orthop Res 15, 213–220, doi:10.1002/jor.1100150209 (1997).

13. Sanchez-Adams, J., Willard, V. P. & Athanasiou, K. A. Regional variation in the mechanical role of knee meniscus glycosaminoglycans. J Appl Physiol (1985) 111, 1590–1596, doi:10.1152/japplphysiol.00848.2011 (2011).

14. Habuchi, H., Yamagata, T., Iwata, H. & Suzuki, S. The occurrence of a wide variety of dermatan sulfate-chondroitin sulfate copolymers in fibrous cartilage. J Biol Chem 248, 6019–6028 (1973).

15. Scanzello, C. R. et al. Fibronectin splice variation in human knee cartilage, meniscus and synovial membrane: observations in osteoarthritic knee. J Orthop Res 33, 556–562, doi:10.1002/jor.22787 (2015).

16. Chockalingam, P. S., Glasson, S. S. & Lohmander, L. S. Tenascin-C levels in synovial fluid are elevated after injury to the human and canine joint and correlate with markers of inflammation and matrix degradation. Osteoarthritis Cartilage 21, 339–345, doi:10.1016/j.joca.2012.10.016 (2013).

17. Ghadially, F. N., Lalonde, J. M. & Wedge, J. H. Ultrastructure of normal and torn menisci of the human knee joint. J Anat 136, 773–791 (1983).

18. Scotti, C., Hirschmann, M. T., Antinolfi, P., Martin, I. & Peretti, G. M. Meniscus repair and regeneration: review on current methods and research potential. European cells & materials 26, 150–170, doi:10.22203/ecm.v026a11 (2013).

19. Fisch, K. M. et al. Identification of transcription factors responsible for dysregulated networks in human osteoarthritis cartilage by global gene expression analysis. Osteoarthritis Cartilage 26, 1531–1538, doi:10.1016/j.joca.2018.07.012 (2018).

20. McCulloch, K., Litherland, G. J. & Rai, T. S. Cellular senescence in osteoarthritis pathology. Aging Cell 16, 210–218, doi:10.1111/acel.12562 (2017).

21. Jeon, O. H. et al. Local clearance of senescent cells attenuates the development of post-traumatic osteoarthritis and creates a pro-regenerative environment. Nat Med 23, 775–781, doi:10.1038/nm.4324 (2017).

22. Sun, H. et al. Single-cell RNA-seq analysis identifies meniscus progenitors and reveals the progression of meniscus degeneration. Annals of the rheumatic diseases 79, 408–417, doi:10.1136/annrheumdis-2019-215926 (2020).

23. Ji, Q. et al. Single-cell RNA-seq analysis reveals the progression of human osteoarthritis. Ann Rheum Dis 78, 100–110, doi:10.1136/annrheumdis-2017-212863 (2019).

24. Gan, Y. et al. Spatially defined single-cell transcriptional profiling characterizes diverse chondrocyte subtypes and nucleus pulposus progenitors in human intervertebral discs. Bone Res 9, 37, doi:10.1038/s41413-021-00163-z (2021).

25. Wu, T. et al. clusterProfiler 4.0: A universal enrichment tool for interpreting omics data. Innovation (N Y) 2, 100141, doi:10.1016/j.xinn.2021.100141 (2021).

26. Hanzelmann, S., Castelo, R. & Guinney, J. GSVA: gene set variation analysis for microarray and RNA-seq data. BMC Bioinformatics 14, 7, doi:10.1186/1471-2105-14-7 (2013).

27. Aibar, S. et al. SCENIC: single-cell regulatory network inference and clustering. Nat Methods 14, 1083–1086, doi:10.1038/nmeth.4463 (2017).

28. Van de Sande, B. et al. A scalable SCENIC workflow for single-cell gene regulatory network analysis. Nat Protoc 15, 2247–2276, doi:10.1038/s41596-020-0336-2 (2020).

29. Kumar, A. et al. Specification and Diversification of Pericytes and Smooth Muscle Cells from Mesenchymoangioblasts. Cell Rep 19, 1902–1916, doi:10.1016/j.celrep.2017.05.019 (2017).

30. Eilken, H. M. et al. Pericytes regulate VEGF-induced endothelial sprouting through VEGFR1. Nat Commun 8, 1574, doi:10.1038/s41467-017-01738-3 (2017).

31. Sugihara, K., Sasaki, S., Uemura, A., Kidoaki, S. & Miura, T. Mechanisms of endothelial cell coverage by pericytes: computational modelling of cell wrapping and in vitro experiments. J R Soc Interface 17, 20190739, doi:10.1098/rsif.2019.0739 (2020).

32. Naba, A. et al. The matrisome: in silico definition and in vivo characterization by proteomics of normal and tumor extracellular matrices. Mol Cell Proteomics 11, M111 014647, doi:10.1074/mcp.M111.014647 (2012).

33. Bouchareb, R. et al. Proteomic Architecture of Valvular Extracellular Matrix: FNDC1 and MXRA5 Are New Biomarkers of Aortic Stenosis. JACC Basic Transl Sci 6, 25–39, doi:10.1016/j.jacbts.2020.11.008 (2021).

34. Hsueh, M. F., Zhang, X., Wellman, S. S., Bolognesi, M. P. & Kraus, V. B. Synergistic Roles of Macrophages and Neutrophils in Osteoarthritis Progression. Arthritis & rheumatology (Hoboken, N.J.) 73, 89–99, doi:10.1002/art.41486 (2021).

35. Alahdal, M. et al. Potential efficacy of dendritic cell immunomodulation in the treatment of osteoarthritis. Rheumatology (Oxford) 60, 507–517, doi:10.1093/rheumatology/keaa745 (2021).

36. Li, Z. et al. Cystatin C Expression is Promoted by VEGFA Blocking, With Inhibitory Effects on Endothelial Cell Angiogenic Functions Including Proliferation, Migration, and Chorioallantoic Membrane Angiogenesis. J Am Heart Assoc 7, e009167, doi:10.1161/JAHA.118.009167 (2018).

37. Walker, A. M. N. et al. Endothelial Insulin Receptors Promote VEGF-A Signaling via ERK1/2 and Sprouting Angiogenesis. Endocrinology 162, doi:10.1210/endocr/bqab104 (2021).

38. Goldring, S. R. & Goldring, M. B. Changes in the osteochondral unit during osteoarthritis: structure, function and cartilage-bone crosstalk. Nat Rev Rheumatol 12, 632–644, doi:10.1038/nrrheum.2016.148 (2016).

39. Xu, L., Ashkenazi, A. & Chaudhuri, A. Duffy antigen/receptor for chemokines (DARC) attenuates angiogenesis by causing senescence in endothelial cells. Angiogenesis 10, 307–318, doi:10.1007/s10456-007-9084-y (2007).

40. Young, M. D. & Behjati, S. SoupX removes ambient RNA contamination from droplet-based single-cell RNA sequencing data. Gigascience 9, doi:10.1093/gigascience/giaa151 (2020).

41. Wolf, F. A., Angerer, P. & Theis, F. J. SCANPY: large-scale single-cell gene expression data analysis. Genome Biol 19, 15, doi:10.1186/s13059-017-1382-0 (2018).

42. Stuart, T. et al. Comprehensive Integration of Single-Cell Data. Cell 177, 1888–1902 e1821, doi:10.1016/j.cell.2019.05.031 (2019).

43. Butler, A., Hoffman, P., Smibert, P., Papalexi, E. & Satija, R. Integrating single-cell transcriptomic data across different conditions, technologies, and species. Nat Biotechnol 36, 411–420, doi:10.1038/nbt.4096 (2018).

44. Aran, D. et al. Reference-based analysis of lung single-cell sequencing reveals a transitional profibrotic macrophage. Nat Immunol 20, 163–172, doi:10.1038/s41590-018-0276-y (2019).

45. McGinnis, C. S., Murrow, L. M. & Gartner, Z. J. DoubletFinder: Doublet Detection in Single-Cell RNA Sequencing Data Using Artificial Nearest Neighbors. Cell Syst 8, 329–337 e324, doi:10.1016/j.cels.2019.03.003 (2019).

46. van den Brink, S. C., et al. Single-cell sequencing reveals dissociation-induced gene expression in tissue subpopulations. Nat Methods 14, 935–936, doi:10.1038/nmeth.4437 (2017).

47. Denisenko, E. et al. Systematic assessment of tissue dissociation and storage biases in single-cell and single-nucleus RNA-seq workflows. Genome Biol 21, 130, doi:10.1186/s13059-020-02048-6 (2020).

48. Korsunsky, I. et al. Fast, sensitive and accurate integration of single-cell data with Harmony. Nat Methods 16, 1289–1296, doi:10.1038/s41592-019-0619-0 (2019).

49. Hua, K. & Zhang, X. A case study on the detailed reproducibility of a Human Cell Atlas project. Quantitative Biology 7, 162–169, doi:10.1007/s40484-018-0164-3 (2019).

50. Baran, Y. et al. MetaCell: analysis of single-cell RNA-seq data using K-nn graph partitions. Genome Biol 20, 206, doi:10.1186/s13059-019-1812-2 (2019).

51. Zou, Z., Hua, K. & Zhang, X. HGC: fast hierarchical clustering for large-scale single-cell data. Bioinformatics, doi:10.1093/bioinformatics/btab420 (2021).

52. Subramanian, A. et al. Gene set enrichment analysis: a knowledge-based approach for interpreting genome-wide expression profiles. Proc Natl Acad Sci U S A 102, 15545–15550, doi:10.1073/pnas.0506580102 (2005).

53. Liberzon, A. et al. The Molecular Signatures Database (MSigDB) hallmark gene set collection. Cell Syst 1, 417–425, doi:10.1016/j.cels.2015.12.004 (2015).

54. Csardi G., N. T. The igraph software package for complex network research. InterJournal Complex Systems, 1695 (2006).

55. Gu, Z., Eils, R. & Schlesner, M. Complex heatmaps reveal patterns and correlations in multidimensional genomic data. Bioinformatics 32, 2847–2849, doi:10.1093/bioinformatics/btw313 (2016).

56. Shao, X. et al. CellTalkDB: a manually curated database of ligand-receptor interactions in humans and mice. Brief Bioinform 22, doi:10.1093/bib/bbaa269 (2021).

